# Malaria molecular surveillance in the Peruvian Amazon with a novel highly multiplexed *Plasmodium falciparum* Ampliseq assay

**DOI:** 10.1101/2021.11.12.21266245

**Authors:** Johanna Helena Kattenberg, Carlos Fernandez-Miñope, Norbert J. van Dijk, Lidia Llacsahuanga Allcca, Pieter Guetens, Hugo O. Valdivia, Jean-Pierre Van geertruyden, Eduard Rovira-Vallbona, Pieter Monsieurs, Christopher Delgado-Ratto, Dionicia Gamboa, Anna Rosanas-Urgell

## Abstract

Molecular surveillance for malaria has great potential to support national malaria control programs (NMCPs). To bridge the gap between research and implementation, several applications (use cases) have been identified to align research, technology development, and public health efforts. For implementation at NMCPs, there is an urgent need for feasible and cost-effective tools.

We designed a new highly-multiplexed deep sequencing assay (Pf AmpliSeq), compatible with benchtop sequencers, allowing for high accuracy sequencing at higher coverage and lower cost than WGS, targeting genomic regions of interest. The novelty of the assay is in its high number of targets multiplexed in one easy workflow, combining population genetic markers with 13 near full-length resistance genes, applicable for many different use cases. We provide a first proof-of-principle for *hrp2* and *hrp3* deletion detection using amplicon sequencing. Initial sequence data processing can be performed automatically, and subsequent variant analysis requires minimal bioinformatic skills using any tabulated data analysis program.

The assay was validated with a retrospective sample collection (n = 254) from the Peruvian Amazon between 2003 and 2018. By combining phenotypic markers and a within-country 28-SNP-barcode, we were able to distinguish different lineages with multiple resistant (*dhfr/dhps/crt/mdr1*) haplotypes and *hrp2* and *hrp3* deletions, increasing in recent years. We found no evidence suggesting the emergence of ART-resistance in Peru. These findings indicate a parasite population under drug pressure, but susceptible to current antimalarials, and demonstrates the added value of a highly multiplexed molecular tool to inform malaria strategies and surveillance systems.

**Importance:** While the power of next generation sequencing technologies to inform and guide malaria control programs has become broadly recognized, integration of genomic data for operational incorporation into malaria surveillance remains a challenge in most malaria endemic countries.

The main obstacles include limited infrastructure and accessibility to high-throughput sequencing facilities and the need for local capacity to run in-country analysis of genomes at a large enough scale to be informative for surveillance. In addition, there is a lack of standardized laboratory protocols and automated analysis pipelines to generate reproducible and timely results useful for relevant stakeholders.

With our standardized laboratory and bioinformatic workflow, malaria genetic surveillance data can be readily generated by surveillance researchers and malaria control programs in endemic countries, increasing ownership and ensuring timely results for informed decision and policy-making.

## Introduction

The Global Technical Strategy for Malaria 2016-2030 identified the use of high-quality surveillance data for decision-making as an essential pillar for malaria elimination (1). In view of current challenges to malaria control (*e*.*g*. spread of drug resistance, changing transmission intensity, risk of malaria importation in malaria-free areas, and RDT-failure), genetic epidemiology is increasingly recognized in its potential to inform national malaria control programs (NMCPs). By aligning multiple scientific fields in support of NMCP priorities, ‘use cases’ were specified creating scenario’s where genetic surveillance can be informative to decision-making (2, 3), and can be used to develop and implement new technologies.

Molecular markers associated with drug resistance are reliable predictors of treatment responses (2, 4, 5), can provide warning for emerging resistance, guide treatment policies (6), and even replace therapeutic efficacy studies in low malaria transmission areas (4, 5, 7, 8). The value of these markers was demonstrated in Southeast Asia where *P. falciparum* resistance against artemisinin (ART) was associated with mutations in the Kelch protein 13 (K13) (9-12), and resistance mutations in this same gene recently emerged in Africa (13).

In addition, monitoring population genetic markers can guide control and elimination strategies by identifying drug resistance gene flow, distinguishing imported from autochthonous cases and estimating the source of reintroduced cases in malaria-free areas (2, 14-16). Parasite genetics can identify underlying patterns of transmission and transmission intensity (17).

The gene products of *P. falciparum hrp2* and *hrp3* are detected by rapid diagnostic tests (RDTs), and gene deletions resulting in false-negative RDT results were first detected in Peru 10 years ago (18). Since then, there have been more reports from many areas (19-22)(23, 24), raising concerns for the viability of HRP2-based RDTs. The WHO recommends assessing prevalence of *hrp2* and *hrp3* mutants among malaria patients and to change case-management strategies when more than 5% of infections contain deletions (25).

Different genetic markers are required to address the forementioned challenges, and are most efficiently targeted in a single genomic tool (3, 26). Whole genome sequencing (WGS) can provide the highest information content per sample, however, implementing WGS for surveillance is constrained by the limited access to high-throughput sequencers and lack of technical and bioinformatic skills in endemic countries. As a result, sequencing is often performed through genomic consortia, delaying the turn-around-time from sample collection to genetic data, and limiting its usefulness to NMCPs. In addition, obtaining high-quality WGS data from low density infections and dried blood spots (DBS) remains challenging. Consequently, it is rare for genomic surveillance to be used beyond research (27-29).

Targeted Next Generation Sequencing (NGS) tools are emerging that combine numerous targets of interest into one workflow and more time and cost-efficient than WGS or traditional PCR-based approaches with amplicons detected by different techniques (10, 30-33). Laboratory protocols for NGS-assays are well-standardized, sequencing is feasible on benchtop sequencers and automated analysis pipelines are available (2, 6), for example the MaRS assay (34) that targets multiple drug resistance markers. While microsatellites have frequently been used for population genetic surveillance as they are not under evolutionary pressure, these markers are not easily translated to NGS due to the short repetitive nature of the variation. Instead, SNP barcodes can be successfully targeted by NGS (35-37), are informative for connectivity between populations (38, 39) and prediction of infection origin (40, 41). The SpotMalaria platform combining drug resistance markers with a SNP barcode was helpful in tracking the rapid spread of resistant parasites in the Greater Mekong Subregion (42).

Currently, there is no multifunctional tool suitable for more than two ‘use cases’. The characterization of *hrp2* and *hrp3* deletions still relies on PCR assays that classify deletions based on failure to amplify targets (23). The few existing tools that combine population markers with drug resistance target short regions around validated resistance SNPs, missing the potential to detect novel resistance associated mutations. Furthermore, many SNP-panels were designed from genomes across the world and lack the resolution to study subtle patterns on a smaller geographical scale. Therefore, we designed a targeted NGS-assay (Pf AmpliSeq) that combines a specifically designed barcode for the target country, 13 full length resistance associated genes, *hrp2* and *hrp3*, and a *ama1* microhaplotype region. The assay is applicable to DBS and adaptable to different settings. The AmpliSeq technology (43) applies a multiplex PCR to simultaneously amplify a high number of targets in a rapid procedure, and allows overlapping amplicons to cover large genes.

We applied the Pf AmpliSeq assay to Peruvian samples for assay validation. *P. falciparum* in Peru makes a good case-study, as malaria elimination is on the regional agenda (44), with countries first targeting *P. falciparum* elimination due to its lower case load than *Plasmodium vivax*, but also in fear of emerging ART-resistance. South America has been a hotspot for chloroquine (CQ) and sulphadoxine-pyrimethamine (SP) resistance evolution (45). More recently, *K13*-mediated ART-resistance emerged in Guyana (46, 47), and K13-independent delayed parasite clearance after ART-treatment was reported in Suriname (46, 48, 49). *P. falciparum* elimination is also threatened by the increasing prevalence and spread of *hrp2* and *hrp3* deletions (21, 23).

Peru reduced malaria case incidence by ≥40% in 2010 (Figure 1), and, after a resurgence in 2016, cases have been decreasing in the past 5 years after introduction of focal screening and treatment (50). Past high levels of CQ and SP-resistance (51-54) led to Peru becoming one of the first countries in the Americas introducing artemisinin combination therapy (ACT) in 2001 (54-56). However, ACT efficacy and drug-resistant markers have not been monitored in Peru since 2009. In 1998-2001 up to 20% of infections in Peru were *hrp2* deleted, increasing to 53% *hrp2* deletions and 37% deletions of both genes in 2012-2014 (18, 19). In this study, we demonstrate the added value of molecular surveillance in Peru with different ‘use cases’ using our Pf AmpliSeq assay and explore the changes of the parasite population in the past two decades.

**Figure 1.**
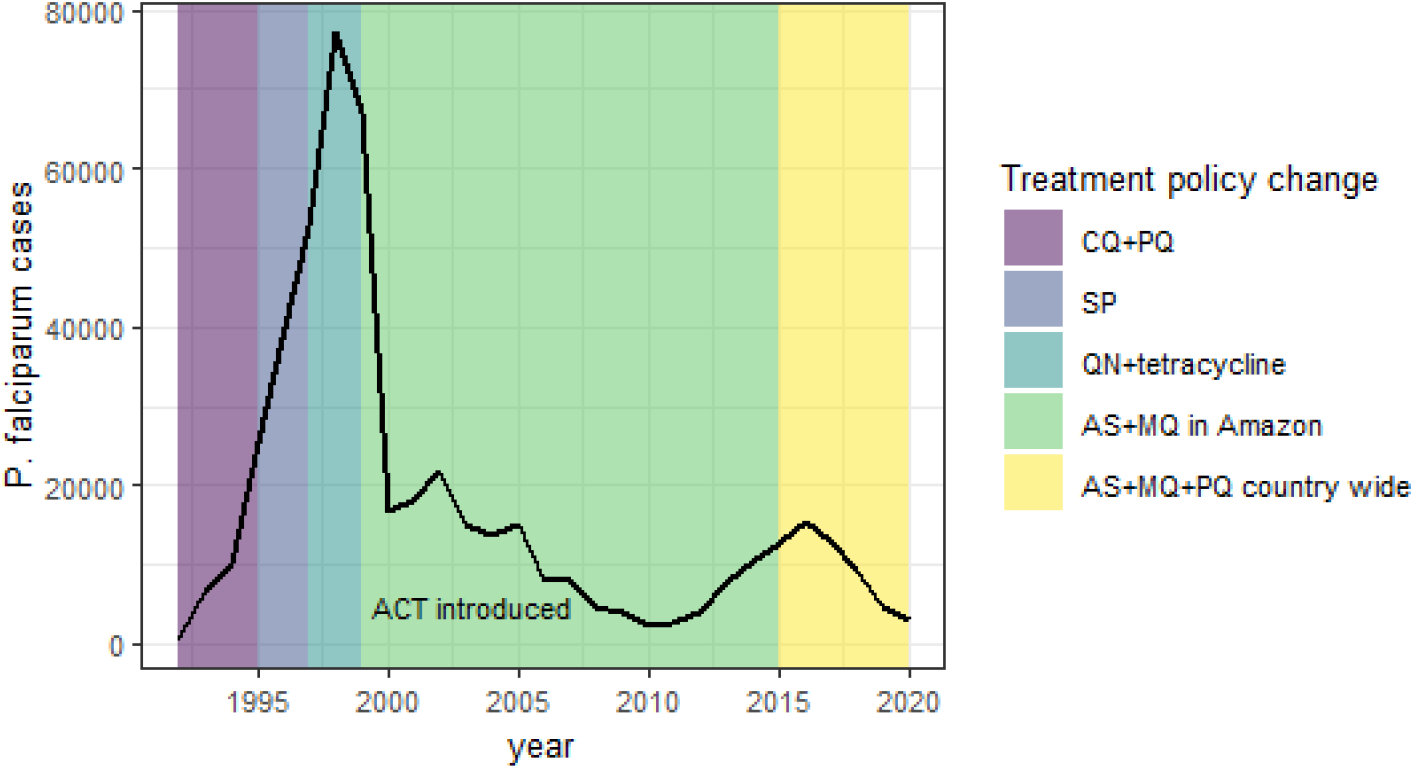
Annual *P. falciparum* cases in Peru. (black line) from 1992-2020, with first-line treatment policy changes (source of data: Centro Nacional de Epidemiología, Prevención y Control de Enfermedades (2020) Sistema de Atención de Solicitudes de Acceso a la Información Pública Vía internet del Ministerio de Salud. Available at: http://www.minsa.gob.pe/portada/transparencia/solicitud/). Artemisinin combination therapies were first introduced in the Amazon region in Peru in 2001, contributing to a dramatic decline in cases. Artesunate mefloquine with primaquine (AS + MQ +PQ) has always been the recommended first-line treatment in the Amazon region (51) with continued high efficacy (57), and AS+SP was used on the North Coast. In 2015, AS+MQ+PQ became the recommended first-line treatment in the whole country (58). Peru reduced malaria case incidence by ≥40% by 2010 using passive case detection (PCD), diagnosis with light microscopy and treatment with AS+MQ as main components of the control program (59). In 2015, with the realization that PCD missed asymptomatic infections (60), focal screening and treatment was introduced, resulting in a further reduction of malaria incidence (50). CQ + PQ = chloroquine + primaquine; SP = sulfadoxine-pyrimethamine; QN = quinine.

## Results

### Pf AmpliSeq design and performance

A panel of 13 *P. falciparum* drug resistant associated genes (Table 1) and corresponding variants of interest, i.e. (putative) resistance associated mutations (Supplementary file 1), were included in the assay design. The selected genes included validated mutations (4, 7) or variants relevant for Peru, i.e. previously reported or associated with historically- and currently-used drugs. Additional targets were: *hrp2* and *hrp3* genes, the apical membrane antigen 1 (*ama1*), conserved *Plasmodium* membrane protein (*cpmp*) (61) and microsatellites (MS) *poly-alpha, ARAII, TA81* and *PfPK2* (62). We designed a barcode of 28 SNPs with in-country resolution in Peru (Supplementary file 3). The final assay included >85% of desired regions in 233 amplicons (Supplementary file 2) with varying amplicon length (55-323 bp). The *cpmp* region and a small section of the *crt* gene failed in the primer design, including crt variant I218F.

**Table 1.**
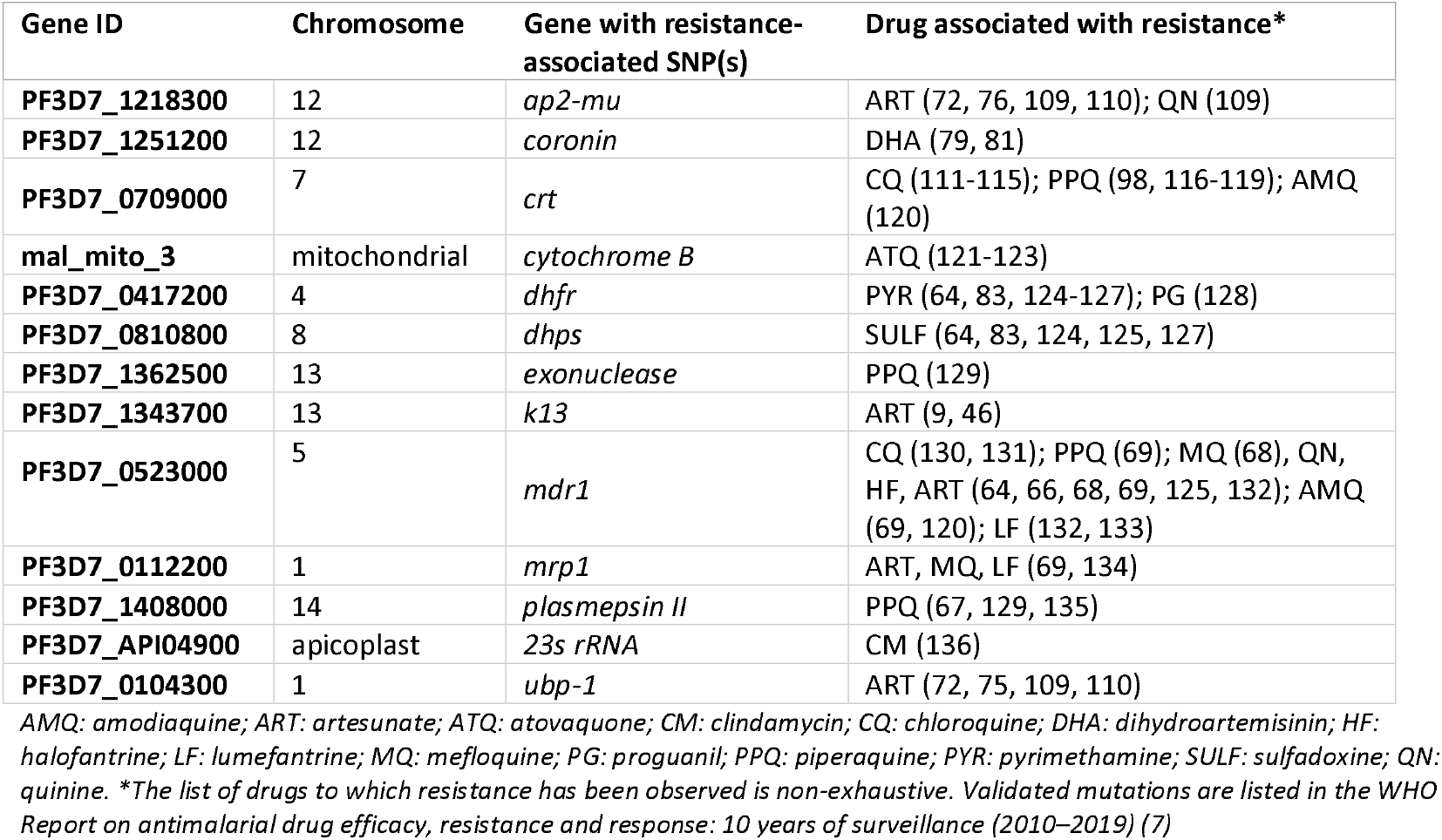
Genes of interest for drug resistance molecular surveillance of malaria in Peru.

We validated the assay with *P. falciparum* cases (n=312, Figure 2) from multiple previous studies in Peru, laboratory strains (n=5), and previously genotyped isolates (n=6) as controls. A high number of reads was generated in the assay (mean 217,037 ± 490,478 paired reads/sample after trimming low quality reads) with a median of 99.6% [range 3.9-99.9%] of trimmed reads aligning to the Pf3D7 genome. Targets regions were covered with a mean 1,336±3,627 paired reads (range 20-43,795). To improve nucleotide diversity in the run (30% GC content), we tested a 20% spike-in of PhiX-library, however, this is not recommended as it did not result in a more high-quality reads (9.7%±10.6 poor quality reads were trimmed without spike-in *vs*. 10.3%±7.3 with spike-in), and reduced total read number.

**Figure 2.**
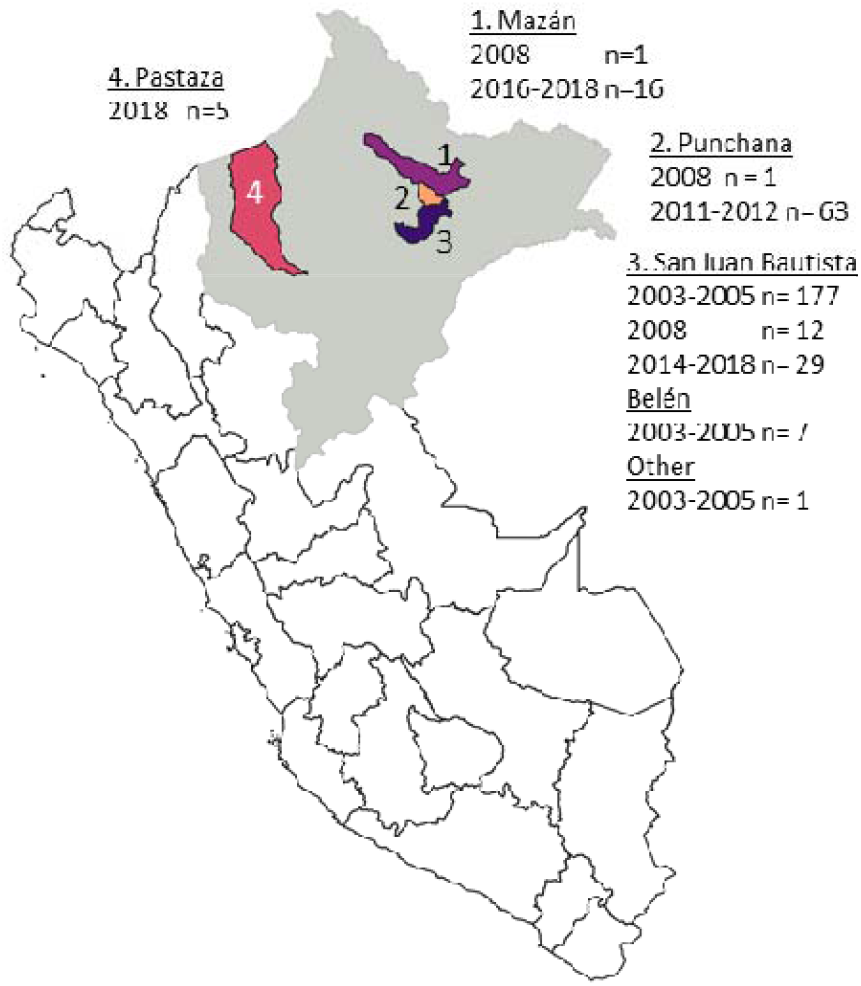
Map of study sites of retrospective sample collection from the Peruvian Amazon region Loreto (grey). The majority of samples were collected in Maynas Province (areas 1, 2, and 3): samples collected in/near the San Juan Bautista district (area 3), which covers part of the urban area of Loreto’s capital Iquitos and peri-urba communities south of Iquitos (n=226), and samples from the Mazán district (area 1; n =17) and Punchana district (area 2; n =64) with remote communities living near forest rivers such as the Amazon River and Napo River, North of Iquitos. A final collection of samples from 2018 was available from the Northern Amazon region in Datem del Marañón Province, Pastaza District (area 4, n=5). *P. falciparum* positive samples from previous studies wer selected based on geographical representation and parasite density (≥100 p/µl by PCR; geometric mean density 4300±3.1 p/µl).

### Depth of coverage

Median amplicon depth of coverage of aligned high quality reads past filter (DP) was 82.9 [IQR 42.9-116.1] (Figure S1). Fifteen (6.4%) amplicons had poor DP (<10; Table S1), while six (2.6%) had high DP (>150; Table S2). Among those, four *hrp2* and *hrp3*-amplicons had lower DP in the study samples, but not in controls, due to high prevalence of deletions in these genes. Lower DP in other amplicons is likely due to sequence variation in primer binding sites, resulting in poor amplification and high amount of missing data in these regions, *e*.*g*. A220S in *crt*. In contrast, two *hrp2*-amplicons had much higher depth than average, possibly due to cross-alignment between *hrp2* and *hrp3* repetitive regions. In 23/233 amplicons no variants (*i*.*e*. only reference alleles) were detected despite good amplification and sequencing (Table S3), and these amplicons were in conserved regions of drug resistance genes.

### Uninfected controls

Primer specificity was tested with four uninfected human blood samples (negative controls), and none of these resulted in genotypes in the assay target region. We did observe background sequences, with a very low number of reads corresponding to multiple unspecific hits to Plasmodium and human sequences; all of them outside the assay target regions and below the filtering threshold.

Eight stored previous library preparations (incl. control samples and one human negative) were included to test replicability, but this resulted in a contamination in the human negative. In the initial run this control was not contaminated, and none of the new library preparations were contaminated with these specific genotypes.

### Parasite density limit and selective whole genome amplification

DP decreased while missingness increased with decreasing parasite density in a dilution of 3D7 (6000 to 6 parasites/µl (p/µl)) at a DNA concentration mimicking dried blood spots (DBS). At the lowest density (6 p/µl), DP was only 10, and 60% of loci were missing. Selective whole genome amplification (sWGA) prior to library preparation improved the number of high quality reads at parasite densities below 60 p/µl (Figure S2); with 10-fold more reads at 60 p/µl to 90-fold at 6 p/µl.

### Error-rate and reproducibility

Sequencing accuracy was higher in biallelic SNPs (mean error-rate 0.008%±0.004) compared to indels (0.02%±0.005) (Table S4). At parasite densities >6 p/µl, sWGA is not recommended, as error-rates were higher with sWGA (0.13% ± 0.06) than without (0.05% ± 0.01), with little gain in coverage.

The reproducibility of the assay was tested, with a median difference of 4 SNPs (0.6% of SNP loci; 0.007% of 57445bp targeted in the assay) between sample and control replicates observed. With indels and multiallelic variants, a median difference of 50 alleles (2.8% of loci; 0.09% of 57445bp) was observed.

In comparison to previous results with laboratory strains and control samples, 97.6% of genotypes were accurate (Table S5). In the control samples, 7.9% additional heterozygous genotypes were detected by AmpliSeq, which might have been previously undetected minor clones due to lower sequencing depth in the SpotMalaria pipeline and WGS data.

### Complexity of Infection

Minority clones could be detected in the Pf AmpliSeq assay up to a 80:20 of 3D7:Dd2 mixture with 43.8% (28/64) Dd2-loci detected (Table S6). Below 95:5 few Dd2-genotypes were detected.

The complexity of infection (COI) estimates the number of genetically distinct clones in an infection, which can be used as a proxy for transmission intensity (16, 63). We applied different methods to determine COI from NGS, but results varied considerably by method (Table S7). Therefore, we determined the most frequent value (mode) of COI, which showed an increasing proportion of multiple clone infections in Peru over time (13.8% in 2003-2005; 28.8% in 2008-2012; 33.3% in 2014-2018; Χ^2^ p=0.0005) (Figure S3).

### Use case *hrp2* and *hrp3* gene deletions

Compared to prior PCR-classification, all samples (10/10) were correctly classified for ‘RDT-failure’ (deletion of both genes) *vs*. ‘RDT-detectable’ (presence of one or both genes) using the read depth of *hrp2* and *hrp3* amplicons. For 6/10 samples, classification was correct for both genes, but in 4 four samples the *hrp2*-classification was inconclusive (Table S8), due to discrepancy in classification between amplicons.

With the Pf AmpliSeq assay, 94.9% (241/254) of study samples could be classified as ‘RDT-failure’ or ‘RDT-detectable’. The proportion of individuals with ‘RDT-failure’ increased from 17.7% in 2003-2005 to 42.6% in 2008-2012 and 73.3% in 2014-2018 (Χ^2^, p>0.001). While *hrp3* presence *vs*. absence could be determined for most samples (96.5%), 26.8% of hrp3+ samples were inconclusive for hrp2 (Figure S9), but since they were not *hrp3* deleted, were classified as ‘RDT-detectable’.

### Barcode performance

All 28 barcode SNPs were detected in the assay (Table S9), although one (Pf3D7_12_1127001) was genotyped as an indel. In the study samples, population minor allele frequencies (MAF) of the barcode SNPs ranged between 0.35-0.50 at 9 loci, between 0.1-0.34 at 12 loci and <0.1 at 7 loci. We did not observe the minor allele of one of the barcode SNPs (Pf3D7_05_921893) in the study samples, although it was detected in laboratory strains (n=5), and 5/6 control samples.

### Use case transmission intensity: Genetic diversity & differentiation

Principal component analysis (PCA) showed sample clustering by year rather than collection site (Figure S4), therefore the samples were divided in three time periods for subsequent analyses: 1) early time period (2003-2005), 2) intermediate time period (2008-2012), and 3) most recent period (2014-2018) where little variability was observed.

Genetic diversity (expected heterozygosity, He) of the parasite population, was lower in 2008-2012 (p=0.003) and 2014-2018 (p=0.00004) than in 2003-2005 (Figure 3A). Total population size was small, as the observed heterozygosity (Hobs) was lower than He (p<0.005), indicating that genetic diversity was lower than expected from a population in Hardy-Weinberg equilibrium. The parasites in 2008-2012 and 2014-2018 showed great genetic differentiation (F_ST_>0.25) compared to parasites in 2003-2005 (Figure 3B). The parasite population changed considerably since 2008, with a less diverse and smaller population.

**Figure 3.**
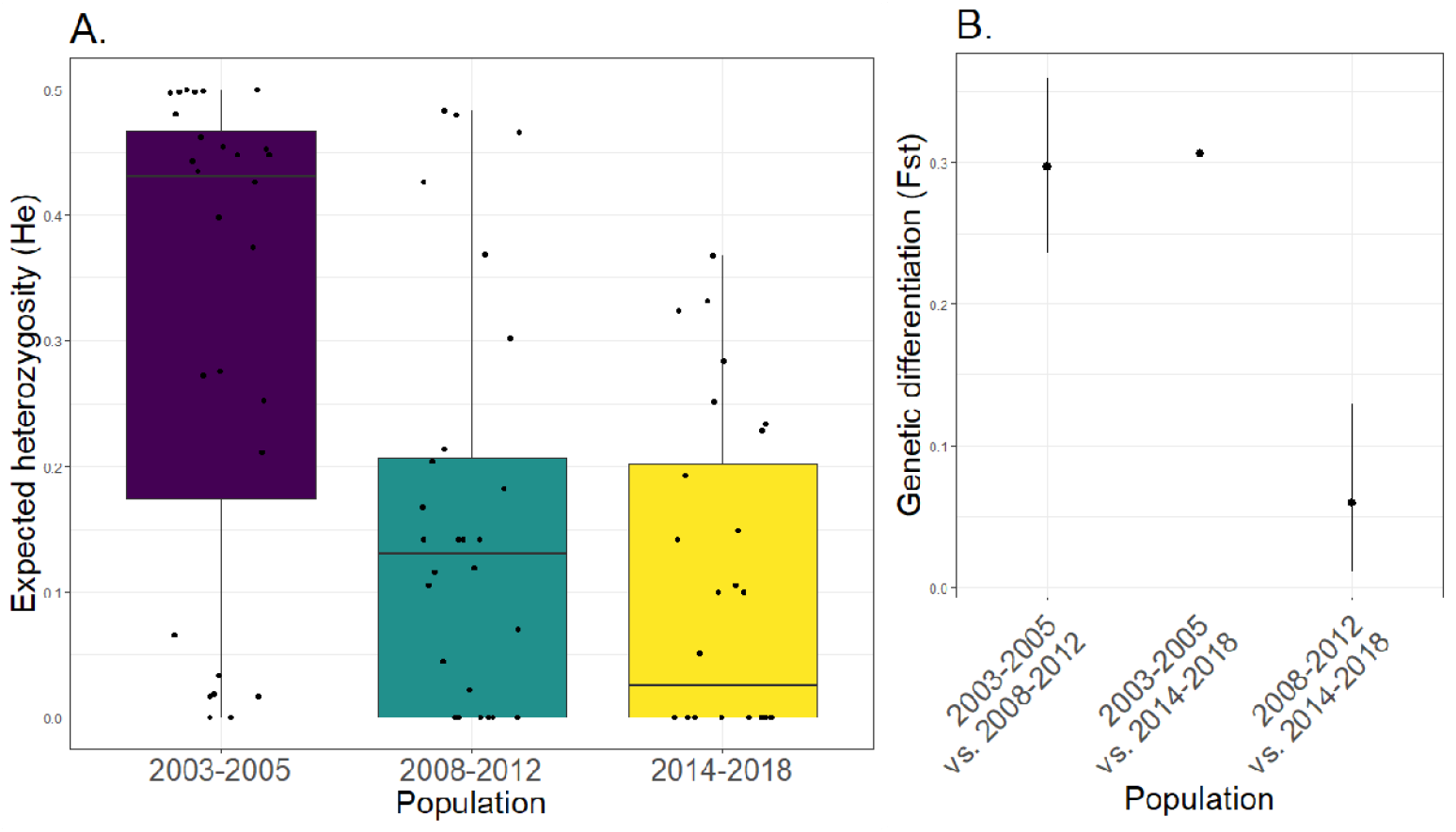
Population genetic statistics. **A)** Genetic diversity (expected heterozygosity, *He*) for each barcode locus (black dots) grouped by time-period. Boxplots show a decrease in the median [interquartile range] of *He* after the first time period (2003-2005). Since differences in He might be due to unequal sampling in districts through time, we assessed He within San Juan Bautista district that was sampled at all three time-periods. This confirmed decreasing diversity over time, with a pronounced decrease in He in 2014-2018 (p >0.001, Figure S5 and table S10). **B)** Genetic differentiation between the three populations measured as F_ST_ (Weir & Cockerham, 1984) using the R package diveRsity. Number of individuals for each population: n_2003-2005_ = 118; n_2008-2012_ = 65; n_2014-2018_ = 38. In Figure S5 He by time and district is plotted and in Figure S6 the differentiation measures G’_ST_ (Hedrick, 2005) and Jost’s D (Jost 2008) are plotted.

### Use case connectivity of parasite populations: Barcode multilocus lineages and population structure

We identified 36 multilocus (ML) lineages with distinct barcodes in the study population over time (Figure 4) and districts (Table 2). The distinct lineages clarify the significant linkage disequilibrium (LD) in the barcode in all time periods and districts (Table S11). In 2003-2005 we observed 23 lineages, and only 11 in 2008-2012, matching the decrease in *He*.

**Table 2.**
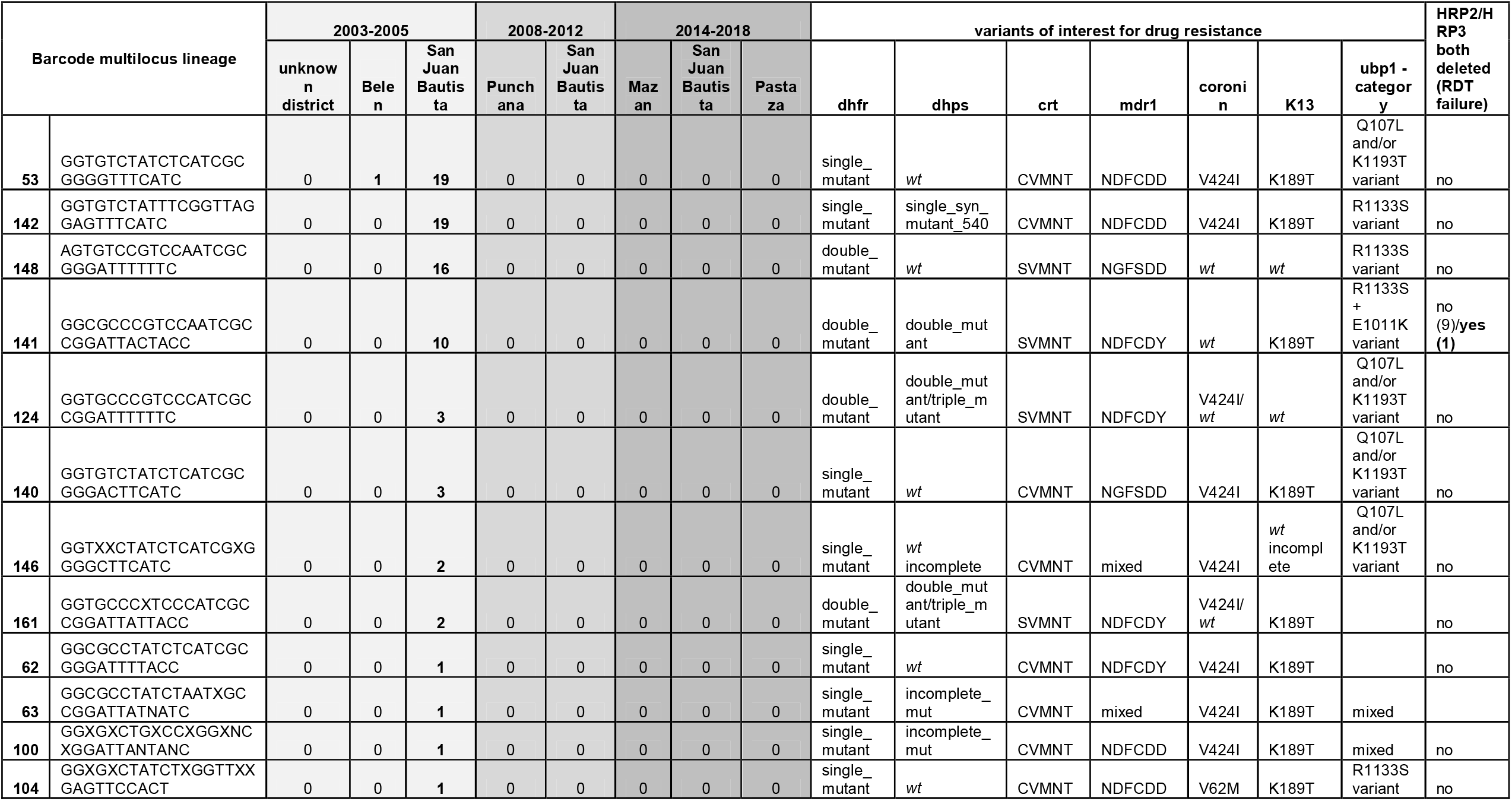
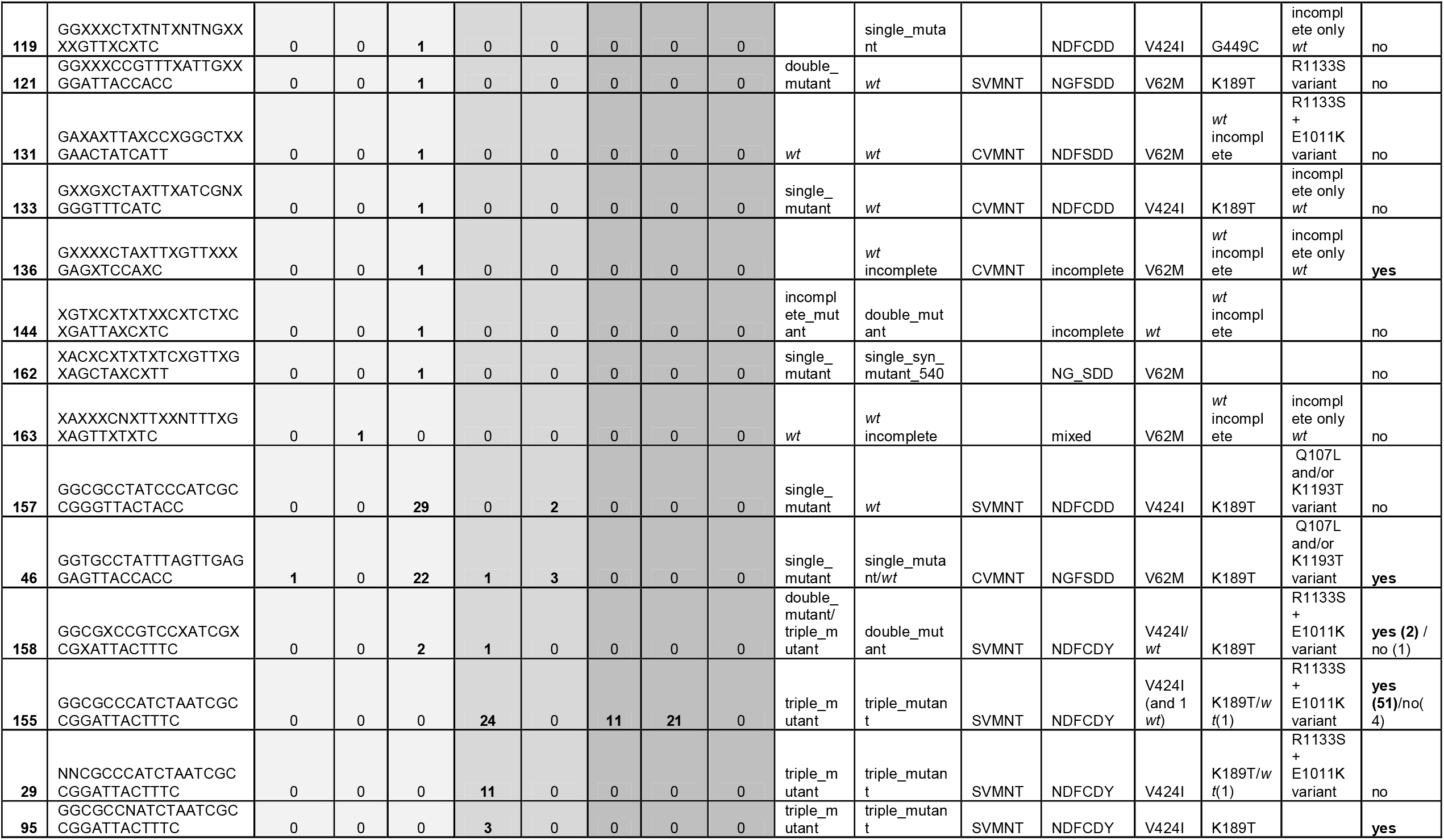
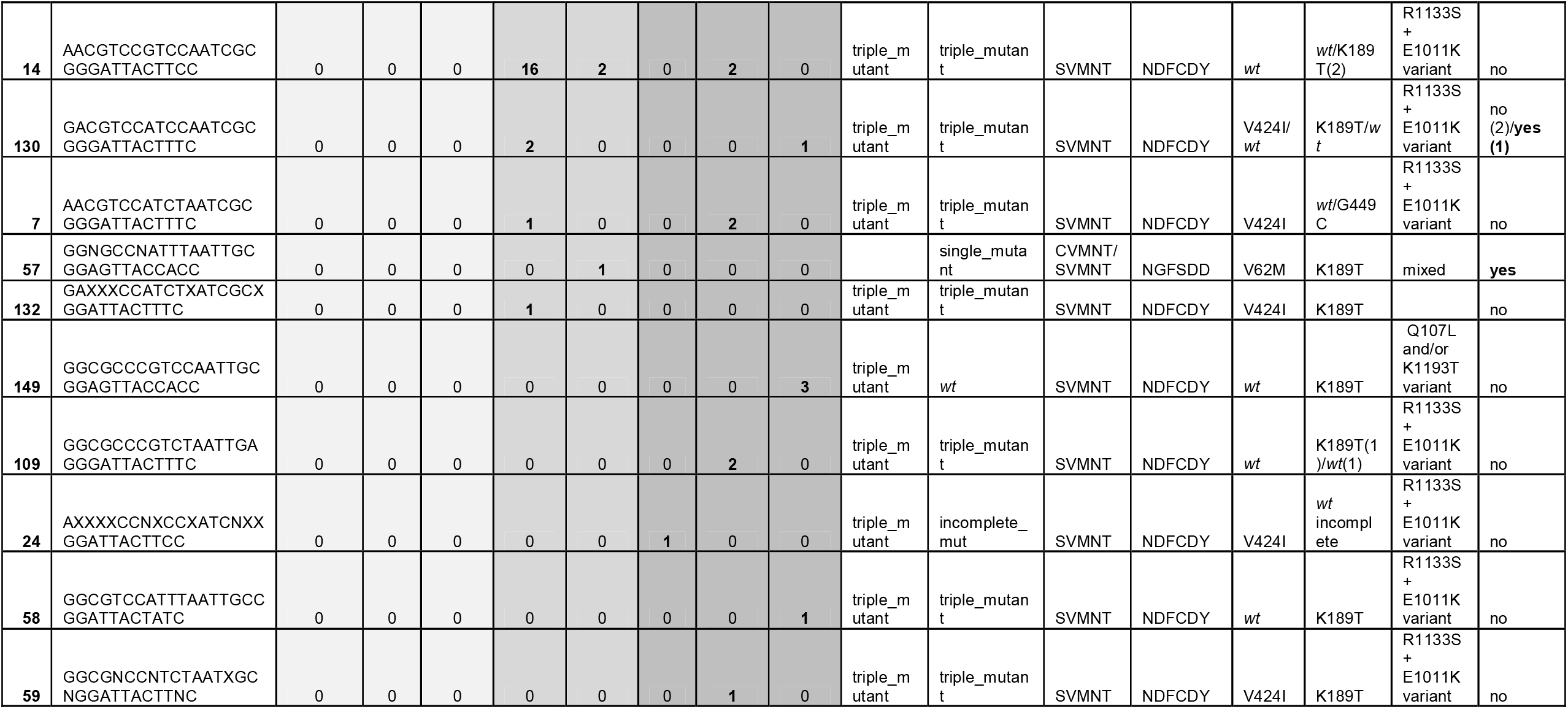
Barcode multilocus lineage characteristics. ML lineages were identified, clustered using genetic distance between barcodes with the poppr package in R, and given a number as identifier. Heterozygote genotypes in the barcode were included, resulting in the creating of some mixed clones barcodes, which include lineages in other lineages. Lineage 29 and 95 are in fact the same as lineage 155, with additional minor clones.

**Figure 4.**
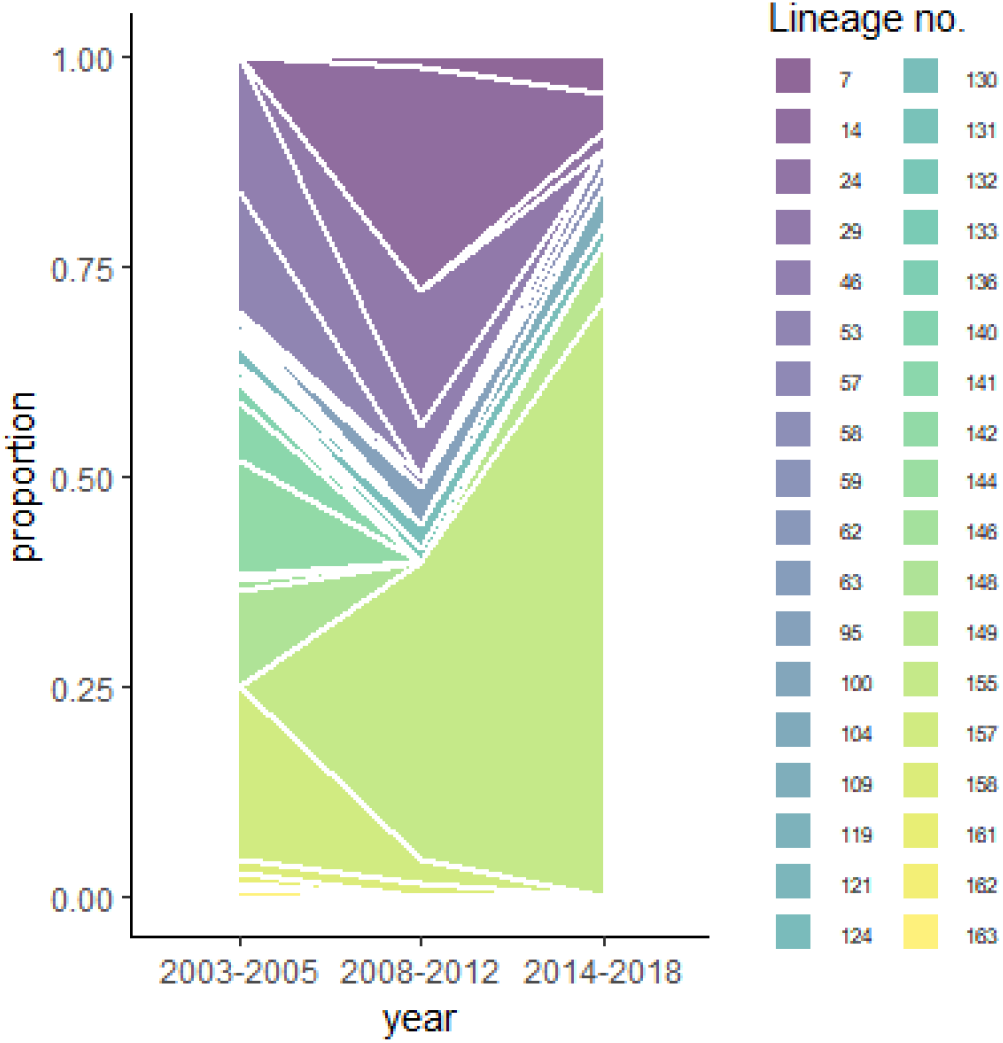
Dynamics of 28-SNP barcode multilocus lineages. Area plot showing the proportions of each ML-lineage detected in the three different time periods in study samples (n=254) from Peru. ML-lineages were identified, clustered using genetic distance between barcodes with the poppr package in R, and given an identifying number. ML-lineages differed by multiple SNPs (≥4 SNPs), enabling lineage classification even when barcodes had some missing or heterozygous genotypes. Lineages are characterized by unique barcodes and resistance genotypes and listed in table 2.

Lineage no. 155 became predominant after 2008, with many related lineages (Figure S7), explaining the small population size and increasing number of barcode alleles that become fixed (2 in 2003-2005; 8 in 2008-2012; 14 in 2014-2018; Table S12). The predominant lineage was in in co-existence with others within district and period. The ML-lineages in later periods (2008-2018) changed considerably since 2003-2005 paralleling observed F_ST_.

With discriminant analysis of principal components (DAPC), the resolution increased to all biallelic variants (nloci=772), and we determined the underlying causes of the population differentiation (Figure 5). The highest contributing alleles along the first axis (2003-2005 vs. 2008-2018) were drug-resistance mutations dhfr C50R and N51I, *mdr1* D1246Y, and *dhps* K540E (Table S13). Contributors to the differentiation along the second axis (2014-2018 and Pastaza) were seven barcode SNPs and *dhfr, mdr1, coronin* and *ubp1*-variants.

**Figure 5.**
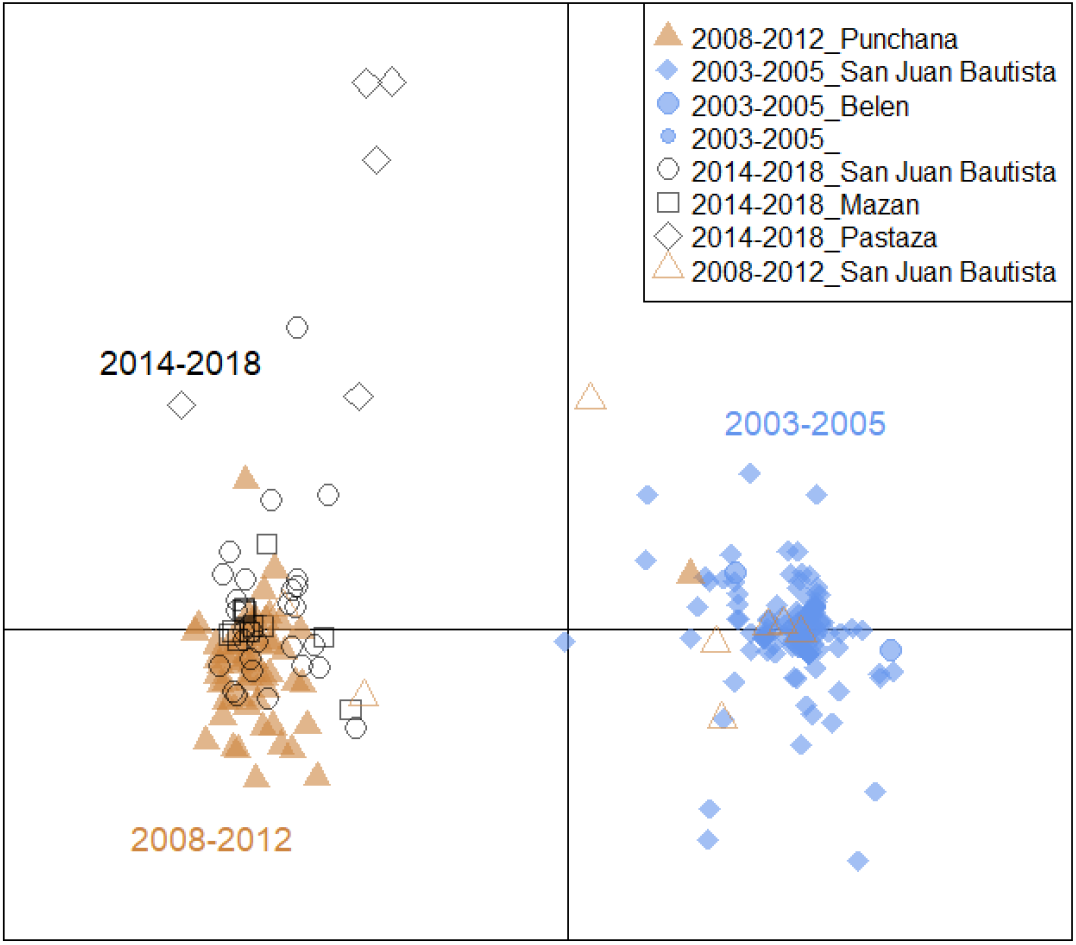
Discriminant analysis of principal components (DAPC). Scatter plot of discriminant analysis (DA) eigenvalues 1 and 2 using all biallelic SNPs in the core targeted region (excl. amplicons targeting repetitive regions and non-nuclear targets), showing a differentiation between Peruvian *P. falciparum* samples collected before 2008 and those afterwards (x-axis, PC1), and a subsequent separation along the y-axis (PC2) from 2014 to more recent years and along geographic distance. SNPs contributing most to the DAPC are listed in Table S13. DAPC was performed with 40 principal components and 7 discriminants as determined through cross-validation. The increased resolution in the DAPC allowed the detection of greater genetic variation in isolates from 2014-2018 (black shapes), compared to lineage analysis.

### Use case drug resistance: Artemisinin resistance-associated genes

The full-length *K13* gene was amplified with 12 overlapping amplicons. We detected several ART-resistance validated SNPs in control samples, but did not observe these SNPs (F446I, N458Y, M476I, Y493H, R539T, I543T, P553L, R561H, P574L and C580Y) in the study samples. We observed several low frequency mutations in the K13-propellor region (V445A, I448K, G449C, V581I, premature stop codon at 613; Table 3), always in mixed infections. Outside the propeller region, the K189T mutation was observed at a high frequency (63%-84%).

**Table 3.** Haplotype frequencies in ART and partner drug resistance associated genes.

|  |  |  | 2003-2005 |  | 2008-2012 |  | 2014-2018 |  | X<sup>2</sup> p-value |
| --- | --- | --- | --- | --- | --- | --- | --- | --- | --- |
|  |  |  | n/n<sub>tot</sub> | % (95%CI) | n/n<sub>tot</sub> | % (95%CI) | n/n<sub>tot</sub> | % (95%CI) |  |
| ART | K13 | G449C* | 1/141 | 0.7% (0.1-4.9) | 0/68 | 0% | 1/45 | 2.2% (0.3-73.4) | 0.03621 |
|  |  | K189T | 94/141 | 66.7% (58.4-74.0) | 42/68 | 61.8% (49.6-73.6) | 25/45 | 55.6% (40.7-73.4) |  |
|  |  | wt | 15/141 | 10.6% (6.5-17.0) | 16/68 | 23.5% (14.9-73.6) | 3/45 | 6.7% (2.1-73.4) |  |
|  |  | wt incomplete | 29/141 | 20.6% (14.6-28.1) | 10/68 | 14.7% (8.0-73.6) | 14/45 | 31.1% (19.2-73.4) |  |
|  |  | missing | 2/141 | 1.4% (0.4-5.6) | 0/68 | 0% | 2/45 | 4.4% (1.1-73.4) |  |
|  | ubp1 | Q107L and/or K1193T type | 60/141 | 42.6% (34.6-50.9) | 6/68 | 8.8% (4.0-18.4) | 3/45 | 6.7% (2.1-19.0) | < 2.2e-16 |
|  |  | R1133S type | 21/141 | 14.9% (9.9-21.8) | 7/68 | 10.3% (4.9-20.2) | 1/45 | 2.2% (0.3-14.6) |  |
|  |  | R1133S + E1011K type | 33/141 | 23.4% (17.1-31.2) | 53/68 | 77.9% (66.4-86.3) | 41/45 | 91.1% (78.3-96.7) |  |
|  |  | mixed Q107L/K1193T+ R1133S | 3/141 | 2.1% (0.7-6.4) | 1/68 | 1.5% (0.2-9.9) | 0/45 | 0 |  |
|  |  | wt or wt incomplete | 24/141 | 17.0% (11.6-24.2) | 1/68 | 1.5% (0.2-9.9) | 0/45 | 0 |  |
|  | coronin | V424I | 78/141 | 55.3% (47.0-63.4) | 41/68 | 60.3% (48.2-71.3) | 37/45 | 82.2% (68.0-91.0) | 0.002743 |
|  |  | V62M | 29/141 | 20.6% (14.6-28.1) | 5/68 | 7.4% (3.1-16.6) | 0/45 | 0% |  |
|  |  | wt | 30/141 | 21.3% (15.3-28.9) | 20/68 | 29.4% (19.7-41.4) | 8/45 | 17.8% (9.0-32.0) |  |
|  |  | mixed | 0/141 | 0% | 1/68 | 1.5% (0.2-9.9) | 0/45 | 0% |  |
|  |  | incomplete | 4/141 | 2.8% (1.1-7.4) | 1/68 | 1.5% (0.2-9.9) | 0/45 | 0% |  |
| SP | dhfr | single mutant (S108N) | 98/141 | 69.5% (61.4-76.6) | 6/68 | 8.8% (4.0-18.4) | 0/45 | 0% | < 2.2e-16 |
|  |  | double mutant + BR + I164L | 32/141 | 22.7% (16.5-30.4) | 0/68 | 0% | 0/45 | 0% |  |
|  |  | triple mutant | 0/141 | 0% | 61/68 | 89.7% (79.8-95.1) | 44/45 | 97.8% (85.4-99.7) |  |
|  |  | incomplete | 3/141 | 2.1% (0.7-6.4) | 0/68 | 0% | 1/45 | 2.2% (0.3-14.6) |  |
|  |  | missing | 6/141 | 4.3% (1.9-9.2) | 1/68 | 1.5% (0.2-9.9) | 0/45 | 0% |  |
|  | dhps | Single mutant K540K | 16/141 | 11.3% (7.0-17.8) | 0/68 | 0% | 0/45 | 0% | < 2.2e-16 |
|  |  | Single mutant A437G | 3/141 | 2.1% (0.7-6.4) | 1/68 | 1.5% (0.2-9.9) | 0/45 | 0% |  |
|  |  | Double mutant | 14/141 | 9.9% (5.9-16.1) | 0/68 | 0% | 0/45 | 0% |  |
|  |  | triple mutant | 2/141 | 1.4% (0.4-5.6) | 48/68 | 70.6% (58.6-80.3) | 30/45 | 66.7% (51.6-79.0) |  |
|  |  | incomplete mutant | 7/141 | 5.0% (2.4-10.1) | 13/68 | 19.1% (11.4-30.4) | 12/45 | 26.7% (15.7-41.6) |  |
|  |  | wt | 80/141 | 56.7% (48.4-64.7) | 4/68 | 5.9% (2.2-14.8) | 2/45 | 4.4% (1.1-16.4) |  |
|  |  | wt incomplete | 19/141 | 13.5% (8.7-20.2) | 2/68 | 2.9% (0.7-11.2) | 1/45 | 2.2% (0.3-14.6) |  |
| CQ and MQ | crt (positions 72-76) | CVMNT | 65/141 | 46.1% (38.0-54.4) | 2/68 | 2.9% (0.7-11.2) | 0/45 | 0% | 8.37E-16 |
|  |  | SVMNT | 49/141 | 34.8% (27.3-43.0) | 60/68 | 88.2% (78.0-94.1) | 32/45 | 71.1% (56.1-82.6) |  |
|  |  | CVMNT/SVMNT mixed | 0/141 | 0% | 1/68 | 1.5% (0.2-9.9) | 0/45 | 0% |  |
|  |  | incomplete | 27/141 | 19.1% (13.4-26.6) | 5/68 | 7.4% (3.1-16.6) | 13/45 | 28.9% (17.4-43.9) |  |
|  | mdr1 (positions 86, 144, 184, 1034,1042, 1246) | NDFCDD | 61/141 | 43.3% (35.3-51.6) | 1/68 | 1.5% (0.2-9.9) | 0/45 | 0% | < 2.2e-16 |
|  |  | ND_CDD | 2/141 | 1.4% (0.4-5.6) | 0/68 | 0% | 0/45 | 0% |  |
|  |  | NDFCDY | 17/141 | 12.1% (7.6-18.6) | 60/68 | 88.2% (78.0-94.1) | 42/45 | 93.3% (81.0-97.9) |  |
|  |  | NGFSDD | 43/141 | 30.5% (23.4-38.6) | 5/68 | 7.4% (3.1-16.6) | 0/45 | 0% |  |
|  |  | NG_SDD | 4/141 | 2.8% (1.1-7.4) | 0/68 | 0% | 0/45 | 0% |  |
|  |  | NDFSDD | 1/141 | 0.7% (0.1-4.9) | 0/68 | 0% | 0/45 | 0% |  |
|  |  | mixed | 5/141 | 3.5% (1.5-8.3) | 0/68 | 0% | 0/45 | 0% |  |
|  |  | incomplete | 8/141 | 5.7% (2.8-11.0) | 2/68 | 2.9% (0.7-11.2) | 3/45 | 6.7% (2.1-19.0) |  |

\*1/2 G449C also accompanied with K189T; both cases mixed inf.
*dhps* double mutant (A437G
+A581G)
*dhps* triple mutant (A437G +A581G + K540E)
wt = wilde type
note: The *ubp1*-haplotype with R1133S and E1011K was frequently observed with additional SNP mutations (K764N, K774N, D777G) and a large insert (EQKY) between amino acid positions 2826/2827.

Forty-four amplicons covered the ubp1 gene (91.8% of the gene), including all variants of interest. Previously reported variants (incl. R3138H) were not detected, but we observed a change in the predominant ubp1-haplotype. In 2003-2005, a haplotype with Q107L, commonly with K1193T, was most frequently observed (42.6%; Table 3). In later years, R1133S-haplotypes (often linked with E1011K) replaced the Q107L-haplotypes.

Eleven amplicons amplified full-length coronin gene. We identified two novel mutations V62M and V424I, and none of the *in vitro* associated variants. V62M was seen at 20.6% in 2003-2005, but decreased over time (Table 3). V424I increased from 55.4% in 2003-2005 to 80.9% in 2014-2018 (Table 3). While V62M is located in the WD-40 beta-propeller domain (containing validated mutations G50E, R100K and E107V), V424I is located just outside this region and is not expected to contribute to ACT resistance.

### Use case drug resistance: CQ and ACT partner drug resistance associated genes

The SP-resistance associated genes were each targeted with 9 amplicons, covering 100% of dfhr and 87% of *dhps*, incl. all variants of interest. In 2003-2005 we observed predominantly single (69.5%) and double (22.7%) *dhfr* mutants, and wildtype (wt) *dhps* (≥56.7%; Table 3). Although SP-use was discontinued in 1999, after 2008, SP-resistance increased with high frequency of triple mutants in *dhfr* and *dhps*. Consistent with previous reports (45, 64), we did not detect *dhf*r C59R in Peru. Therefore, the ‘sextuple’ mutant here is different from the African sextuple mutant, with C59R instead of C50R, but similar super resistant phenotype (65).

Sixteen amplicons covered the *crt* gene (91.1%), missing one variant of interest (I218F) associated with PPQ resistance. CQ resistance (CQR) crt haplotypes, CVMNT (CQR) and SVMNT (highly CQR), increased over time (Table 3) with 100% SVMNT in 2014-2018. The mutation I356V, whose contribution to CQR is unclear, was frequent (>93%) in all time periods.

Eighteen amplicons covered the *mdr1* gene (87%), targeting all variants of interest associated with CQ and MQ-resistance (7). While the *mdr1* haplotypes NDFCDD and NGFSDD were predominant in 2003-2005, in later years it was mainly the NDFCDY haplotype (Table 3). There are conflicting results on the association of NDFCDY with susceptibility to MQ, AS and CQ (64, 66). The *mdr1* D144G SNP has so far only been reported in Peru (annotated as D142G in (64)) and is decreasing in recent years.

Copy number variations (CNVs) in *mdr1* and *plasmepsin II* associated respectively with PPQ (67) and MQ (68) resistance were not previously observed in Peru (69). We confirmed single copies of both genes by qPCR in a subset of 78 samples (Figure S8). While both genes were sequenced in our assay, the lack of increased copy numbers resulted in insufficient data to validate CNV detection by Pf AmpliSeq.

We did not detect many variants of interest in the other targeted genes, with 10.6% of the S160N mutant in 2003-2005 in the ap2-mu gene (9 amplicons covering 99.1% of the gene), which decreased in later periods (1.5% and 2.2%). No variants of interest were observed in *pfmrp1* (95.1% covered), *pfexonuclease* (98.6% covered), *cytochrome B* (100% covered) and *23S rRNA* gene (100% covered).

### Use case drug resistance gene flow: drug resistance and parasite lineage evolution

Between 2003 and 2005, we observed lineages with predominantly single and double *dhfr* mutants, *wt dhps*, CVMNT and SVMNT crt-haplotypes, multiple *mdr1-*haplotypes and presence of *hrp2* and *hrp3* deletions (Table 2). From 2008, the majority of parasite lineages harbored sextuple *dhfr/dhps* mutants, crt SVMNT, mdr1 NDFCDY, and *hrp2* and/or *hrp3-*deletions. Parasites with this *dhps/dhfr/crt/mdr1* resistance haplotype and both *hrp2* and *hrp3* deleted were first reported in few cases from Peru in 2006 (64) and called the BV_1_-lineage. Here, we observed many parasites with the same ‘BV_1_-type’ resistance haplotype, although not always with *hrp2* and *hrp3-*deletions. With the greater resolution in the Pf AmpliSeq assay, we observed many related lineages with this resistant haplotype that differ in the barcode genotype, presence of *hrp2* and *hrp3-*deletions, and coronin SNPs (Table 2). This indicates gene exchange between lineages contrasting the idea of isolated clonal lineages in Peru.

The BV_1_-type lineages are closest related to earlier lineages 158, 141 and 63 in our study samples (Figure S7), but the triple *dhfr* mutants are not accompanied by the secondary mutations (I164L and the “Bolivian Repeat” (BR), a silent 5 amino acid insertion before codon 30), indicating that they did not emerge from the double *dhfr* lineages circulating in earlier years. In conclusion, the combination of different markers in the Pf AmpliSeq assay allowed the identification of multiple recombining *P. falciparum* lineages in Peru that share increasing SP and CQ resistance, as well as RDT failure, in more recent years.

## Discussion

We designed a new highly-multiplexed sequencing assay (Pf AmpliSeq) targeting 13 antimalarial resistance genes, combined with a country-specific 28-SNP barcode for population genetic analysis in Peru and *hrp2* and *hrp3* genes to detect deletions that can cause false-negative RDT results. The novelty and strength of this assay is in the many different types of markers multiplexed in one easy workflow, making it suitable for many surveillance ‘use cases’. Compared to other amplicon sequencing assays (30, 34, 70) and genetic surveillance platforms (71), the Pf AmpliSeq includes the highest number of drug resistance associated genes, combined with an in-country SNP-barcode, as well as diagnostics resistance markers.

This assay performs well on DBS samples with parasite densities ≥60 p/µl, and allows for high accuracy sequencing at higher depth of coverage (100-1000 fold vs. 50-100 fold) and lower cost (Table S14) than WGS. At densities <60 p/µl, sWGA prior to the Pf AmpliSeq assay increases the number of reads, but also the error rate and cost. Initial raw data processing can be performed automatically using software of the sequencer, with no need for linux-based pipelines as we have used (giving us more flexibility and insight during the validation). Subsequent variant analysis can be performed on a regular computer with minimal bioinformatic skills using any software that can analyze tabulated data.

While the assay was capable at detecting ART-resistance validated SNPs in *K13*, only non-validated SNPs were detected at low frequency in the Peruvian samples used for assay validation. Without presence of validated markers in *K13*, there is no evidence suggesting the emergence of ART resistance in Peru in recent years (2003-2018). Mutations in other genes in the endocytosis pathway have been associated with decreased sensitivity to ART in *in vitro* studies and changes in *ubp1-*haplotypes paralleled the emergence of *K13* markers in Thailand (72-81). However, the observed *ubp1* and *coronin* mutations here are uncharacterized. Genetic variants in KIC7, Eps15, Formin2 associated with *in vitro* ART resistance (74-76, 78, 82) were not reported yet when the assay design was being completed. These genes will be included in future versions of the assay.

This is the first proof-of-principle of an NGS-amplicon assay targeting *hrp2* and *hrp3-*deletions, detecting 56.3% samples with both *hrp2* and *hrp3* deleted. This might be an underestimation, since infections containing mixed strains with and without deletions might be classified as ‘RDT-detectable’. It was difficult to classify *hrp2* for 22% of samples, due to conflicting results between *hrp2* amplicons. The difference in read depth with different *hrp2* amplicons suggests that potentially only a part of the gene is deleted, with (part of) the first exon still present. Better characterization of the genomic structure of *hrp2-*deletions could improve amplicon design. In addition, copy number calling tools could be explored for more systematic classification of *hrp2* and *hrp3-*deletions.

With the combination of phenotypic markers and the 28-SNP-barcode in the Pf AmpliSeq assay, we were able to detect changing allele frequencies in resistance associated genes, but also to identify temporal genetic differentiation in the parasite population, alongside a decrease in genetic diversity. The barcode served to distinguish different lineages in multiple resistant (*dhfr/dhps/crt/mdr1*) haplotypes circulating in recent years, and enabled the investigation of drug resistance evolution.

SP- and CQ-resistance in South America was reportedly spread from a single origin in the lower Amazon (45), with five distinct clonal lineages circulating in Peru before 2006 (19, 64, 83), followed by frequent outcrossing and lineage evolution (83). With the Pf AmpliSeq, we observe the same SP/CQ-resistant lineages and their evolution over time, with increased resolution and additional variants detected in other genes.

The BV_1_-type lineages that increasingly dominate the Peruvian parasite population after 2008, are highly SP/CQ-resistant, and can frequently escape HRP2-based RDTs. The Pf AmpliSeq assay could characterize outbreaks, such as the BV_1_-outbreaks in Tumbes (84) and Cusco (85). What pressure selected the BV_1_- type lineages is unclear, as there is no clear evidence of ART-MQ resistance (absence of *K13*-variants and *mdr1* CNV) and HRP2-based RDTs are not commonly used in Peru. The high level of CQR (in *crt* and *mdr1*) could be the result of CQ-treatment of co-endemic *P. vivax* cases. Co-infections are frequent (86), and can occur with low density *P. falciparum*, and frequently misdiagnosed and treated as P. vivax mono-infection (83). When the BV_1_-type lineages first appear (2011), we observed strong population differentiation and loss of secondary *dhfr* mutations, which supports the suggestions from others that this lineage was introduced from other parts of the Amazon (84, 87), possibly Colombia (20, 21) or Suriname (49). Analysis of isolates from neighboring countries could shed light on the evolution and spread of resistance and *hrp2* and *hrp3* deletions in the region. Predicting the origin of imported infections based on SNPs is possible (40, 41, 88), however, samples from neighboring countries were not available to investigate the spatial dynamics within this study. Existing public WGS datasets lack isolates from sufficient countries in South America for a comprehensive analysis.

Ideally, to update genomic regions of interest, periodic WGS of a subset of samples should be performed to investigate the emergence of mutations not targeted by the Pf AmpliSeq assay. Newly identified regions could be added to the design. Similarly, we can exchange the SNP-barcode for application in different countries, resulting in a modular design adaptable to the needs of a particular setting. The assay in its current form is limited to South America. However, we are validating Pf AmpliSeq assays for South East Asia and Sub-Saharan Africa, including also other ART-resistance genes, and *P. vivax* Ampliseq assays.

Conflicting COI results were obtained due to differences between methods. In addition, the variant calling tool used here assumes a diploid genotype with balanced within-sample allele frequency, which is frequently not the case in malaria complex infections. Alternative haplotype-based analysis approaches are available (e.g. SeekDeep (89), HaplotypR (61), and DADA2 (90)) that might have a greater capability at unraveling complex infections, and more suitable for relatedness analysis (*e*.*g*. identity-by-descent (91)). These methods could work with the existing design, or by adding microhaplotypes (70). However, they are less easy to use for drug resistant markers and overlapping amplicons.

As half of the barcode SNPs became fixed in the most recent years, the usefulness of the barcode in future years might be limited. This likely reflects a true decrease in diversity as the sample size was sufficient, and there are closely related clonal lineages. The sample size in this study was comparable to earlier reports from Peru (e.g. 220 samples in 1999 (83), 104 samples from 1999 and 62 from 2006-2007 (64)), but represents a limited proportion of the Peruvian *P. falciparum* population. A more systematic sampling approach of dried blood spots at many more sites within the country would be ideal for routine genetic surveillance.

## Concluding remarks

This study described a *P. falciparum* population in Peru with increasing accumulation of drug and diagnostic resistance and spread of these lineages can be a serious threat to *P. falciparum* elimination throughout the Amazon region. This study highlights the importance of systematic resistance monitoring in the country, which is currently lacking, and offers a surveillance tool for multiple genetic ‘use cases’. Implementation will require a structured approach that will most importantly not unnecessarily burden existing health systems and is sustainable for the long term. A molecular surveillance network (GENMAL) has been established to stimulate wide-scale implementation and standardization of markers for surveillance. This network will be a platform for coordination of research in Peru, data sharing and efficient translation of results into policy-advice and practical guidelines for malaria elimination in Peru. The type of multiplex assay as presented here could be more broadly applicable for the wider Amazon area and South America to fill the void of genetic data from this region.

## Materials and methods

### Study sites, samples and controls

*P. falciparum* positive samples (n=312) from previous studies were selected based on geographical representation (Figure 2) and parasite density (≥100 p/µl by PCR). Samples from 2003-2017 (n=269) were from studies led by Universidad Peruana Cayetano Heredia (UPCH) ((18, 92) and ongoing), and the U.S. Naval Medical Research Unit 6 (NAMRU-6) (n=63). Samples from 2018 (n=5) were the only *P. falciparum* positive samples from a Zero Malaria Programme collection in Loreto in collaboration with UPCH.

DNA was extracted from DBS using the E.Z.N.A Blood DNA Mini Kit (Omega Bio-tek, Georgia, USA) following instructions. Per sample, two pieces of ∼0.5 cm^2^ were cut out and 100μL DNA was eluted. *P. falciparum* parasitaemia was qPCR quantified (93).

The studies were approved by local ethical review boards. Protocols were registered in the Decentralized System of Information and Follow-up to Research (SIDISI) (numbers: 52707, 61703, 101645, 66235, 102725), and one clinical trial registered at clinicaltrials.gov (NCT00373607). The NAMRU-6 study was approved by its Institutional Review Board in compliance with all applicable federal regulations governing the protection of human subjects (protocol NMRCD.2007.0004). Individuals were included in this study only if signed informed consent included a future-use clause, and secondary-use was approved through the Institutional Review Board of the Institute of Tropical Medicine Antwerp (reference 1417/20).

For the final analysis we selected 254/312 (81%) samples with good quality data (<50% missing genotype calls, mean coverage >15) and retaining only one library of replicates (with lowest missingness). With a sample size of 96 samples a MAF of 0.01 with 0.02 (2%) precision could be detected, and with 381 samples MAF=0.01 with (1%) precision (https://epitools.ausvet.com.au/oneproportion).

Laboratory isolates and uninfected human blood were included as controls (Table S15). Control samples (n=6) with known variants from a previous study in Vietnam (94) genotyped with the SpotMalaria pipeline and WGS were also included. DNA of controls (100μL) was extracted using QIAamp DNA Mini Kit (Qiagen) following instructions. sWGA was performed as previously described (95). Genotypes of the laboratory strains were from (69, 96-98).

### SNP-barcode design

A barcode of 28 biallelic SNPs was designed (Supplementary file 3). Briefly, MalariaGEN *Plasmodium falciparum* Community Project data (99) was used to select SNPs (0.35≤MAF≤0.5) contributing to between-country differentiation using DAPC (100). Two SNPs per chromosome were selected with priority for synonymous SNPs with low pairwise-LD.

### Pf AmpliSeq workflow and sequencing

Library preparation was performed using AmpliSeq Library PLUS for Illumina kit (Illumina), AmpliSeq Custom Panels (Supplementary file 2) and AmpliSeq CD Indexes (Illumina) as per the manufacturer’s instructions. DNA concentration for controls and 25 samples was determined using Qubit v3 High sensitivity DNA kit (Invitrogen) and controls were diluted to 1 ng input DNA to mimic DBS samples. Study samples (mean DNA concentration 6.1±0.3 ng/μl) were not diluted. Target regions were amplified with adjusted cycling conditions (1×99⁰C-2min; 21×99⁰C-15sec; 1×60⁰C-8 min) in two reactions, and subsequently combined for final library preparation according to the guidelines. Libraries were quantified using KAPA Library Quantification Kit for Illumina Platforms (KAPA Biosystems), diluted to 2nM with low Tris-EDTA buffer, and pooled. Denatured library pool (18pM) was loaded on a Miseq system (Illumina) for 2×300 paired-end sequencing (Miseq Reagent Kit v3, Illumina). A detailed protocol is available at bio-protocol.org/###. 20% PhiX spike-in (Illumina) was used in one sequencing run to determine the effect on sequencing quality and trade-off in coverage. Generated demultiplexed FASTQ files were processed with an in-house analysis pipeline on a Unix operating system computer (Supplementary file 3).

### Detection of *mdr1* and *pm2* copy number variations by PCR

CNVs in *pm2* and *mdr1* were determined in a subset of samples (n=78) from all time periods and districts using qPCR (94). Samples and controls were tested in triplicate with 20% of samples re-tested for reproducibility with 100% of results in agreement.

### Analysis of Complexity of infection

COI was estimated using the Real McCOIL categorical and proportional methods (101) in R using subsets of variants: 1) all biallelic SNPs, 2) 25 barcode SNPs, 3) “core variants”: biallelic variants excluding repetitive regions (i.e. *hrp2, hrp3* and MS) and mitochondrial and apicoplast regions. Default settings were used for a fixed error, and for the categorical method an upper bound of 10, and initial COI of 5 and for the proportional method an upper bound of 15, and initial COI of 7. We allowed 50% missingness for biallelic SNPs, 20% for the barcode and 40% for core variants.

We also estimated the proportions of single and multiple clone infections based on the number of heterozygous genotypes in 1) the 28-SNP barcode, 2) biallelic SNPs in ama1, and 3) core variants. Samples with ≥1 heterozygous genotype were considered to contain multiple clones (COI≥2). Finally, we estimated COI from the MS amplified regions. As the different methods generated diverging results, the mode (most frequent value of single vs. multiple clone) was determined as final measure for each sample.

### Validation of detection of *hrp2* and *hrp3* deletions

The Pf AmpliSeq performance for *hrp2* and *hrp3* deletion detection was assessed in 10 previously-typed samples (31). The presence or absence of both genes was determined using the mean read depth of *hrp2* and *hrp3* amplicons (Supplementary file 3). Due to the repetitive nature and homologies between *hrp2* and *hrp3* genes we used a conservative cut-off value, resulting in a “grey zone” where deletion/presence was left inconclusive when the majority of amplicons were not in accordance. A final variable for ‘RDT-failure’ (both *hrp2* and *hrp3* absent) vs. ‘RDT-detectable’ (*hrp2* and/or *hrp3* present) was created, allowing the classification of samples that were inconclusive in one of the two genes if the other gene was present.

### Population genetic analysis

Population genetic analyses were performed using the 28-SNP barcode, excluding samples missing >7 SNPs (25%). Samples with heterozygous genotypes were included, justified by little genetic differentiation (F_ST_=0.05) between samples with single clone barcodes vs. samples with heterozygous barcode genotypes. Genetic differentiation was measured as F_ST_ (102), G’ST (103) and Jost’s D (104) using 1000 bootstraps with the R package DiveRsity (105). PCA was performed on the genotype matrix using ‘prcomp’ function (R stats package v4.0.5) using core variants. Prior to PCA, missing genotypes were replaced by the mean allelic frequency at a locus in all samples. Expected heterozygosity (He) was calculated (adegenet package (106, 107) in R) using diploid barcode genotypes. Wilcoxon signed-rank test with continuity correction was used to compare mean observed heterozygosity (Hobs) to He. LD was measured as the standardized index of association 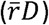 in multilocus analysis using 999 resamplings (method: permutation of alleles) using the poppr package v2.8.6. (108) in R. ML-lineages were defined by grouping isolates with similar barcodes using the Hamming distance and clustered based on the maximum distance (farthest neighbor) using poppr. DAPC (100) was performed with cross-validation, and associated allele loadings for the first four components were determined.

### Analysis of genetic variants

Barcode allele frequencies were calculated from allele depths to reflect population allele frequencies in complex infections (Supplementary file 3). Haplotypes were created by combining genotypes of variants of interest (Supplementary file 1). Frequency tables with confidence intervals were created with the freqtables R-package.

## Data availability

Sample meta data and drug resistant and *hrp2* and *hrp3* haplotypes, barcodes, lineages location, date, etc. is accessible at https://microreact.org/project/aV7RHNCBmG3sJ2rwzchE4k-peru-2003-2018-molecular-surveillance-with-p-falciparum-ampliseq-assay. Raw data (fastq), variant files (vcf), and scripts are available on request. All other data is included in the manuscript and supporting files.

## Acknowledgements

We wish to thank all clinical, microscopy and field staff who supported the sample collections and all participants for making their material available for malaria studies. We acknowledge the support of the Dirección Regional de Salud de Loreto for the study authorization and sample collection activities in Loreto. The following reagents were obtained through BEI Resources, NIAID, NIH: *P. falciparum*, Strain Dd2, MRA-150, contributed by David Walliker; *P. falciparum*, Strain Dd2_R539T, MRA-1255, and *P. falciparum*, Strain CamWT_C580Y, MRA-1251, contributed by David A. Fidock; and *P. falciparum*, Strain IPC 4912, MRA-1241, contributed by Didier Ménard.

## Copyright statement

Some authors of this manuscript are military service members and employees of the U.S. Government. This work was prepared as part of their official duties. Title 17 U.S.C. §105 provides that “Copyright protection under this Title is not available for any work of the United States Government”. Title 17 U.S.C. §101 defines a U.S. Government work as a work prepared by a military service member or employee of the U.S. Government as part of that person’s official duties. The views expressed in this article are those of the authors and do not necessarily reflect the official policy or position of the Department of the Navy, Department of Defense, nor the U.S. Government.

## Author contributions (in alphabetical order per item)

Conception of ideas for the study: ARU, DG, JHK; Selection of targets and design of the assay: ERV, JHK, NvD; Sample collections and coordination of field work: CDG, CFM, DG, HOV, JPvG; Laboratory experiments: CFM, JHK, NvD, PG; Bioinformatics & Data analysis: JHK, LLA, NvD, PM; Writing first draft manuscript: ARU, JHK, CFM, NvD. All authors reviewed and contributed to the final version of the manuscript.

## List of supplementary material

### Supplementary files

**Supplementary file 1: Variants of interest for drug resistance (.csv)**

**Supplementary file 2: PF AmpliSeq design: primer sequences and locations on the genome (.xlsx)**

**Supplementary file 3: Supplementary methods (.docx)**

**Supplementary file 4: Pairwise linkage disequilibrium (rbarD) and allele loadings for within-country SNP selection for *P. falciparum* Ampliseq Peru (.xlsx)**

### Supplementary figures and tables

**Figure S1. Distribution of depth of coverage of aligned high quality reads past filter (format field DP) per amplicon region**.

**Table S1. Amplicons with low genotype depth**.

**Table S2. Amplicons with high genotype depth of coverage (>150)**.

**Table S3. Amplicons in conserved regions**.

**Figure S2. Effect of selective whole genome amplification (sWGA)**

**Table S4. Error rates in 3D7 replicates without and with sWGA in different subsets of loci**.

**Table S5. Genotyping known variants in previously genotyped controls**

**Table S6. Detection limit of variant loci in 3D7-Dd2 mock samples**

**Table S7. Complexity of infection analyses**.

**Figure S3. Proportion multiple clone infections in Peru**.

**Table S8. Pairwise comparison of *hrp2/hrp3* classification by PCR and Pf AmpliSeq**

**Table S9 Barcode SNPs**.

**Figure S4. Principal component analysis of samples (n=254) collected in Peru between 2003 and 2018**.

**Figure S5. Expected Heterozygosity by time period and district**.

**Table S10. p-values for pairwise comparisons of *He***.

**Figure S6. Genetic differentiation between parasite populations in the three time periods**

**Figure S7. Minimum spanning network (nLV graph) of multilocus lineages**

**Table S11. Linkage disequilibrium**

**Table S12. 28-SNP barcode loci that become fixed over time**

**Table S13 Contributions of alleles to DAPC**.

**Figure S8. Copy number variations in A) plasmepsin II gene (pm2) and B) multidrug resistance gene 1 (mdr1)**

**Table S14 Cost comparison AmpliSeq vs WGS**

**Table S15: Laboratory strains included in assay validation**

**Figure S9. Distributions of log mean depth ratio’s for all samples for each *hrp3* amplicon and *hrp2* amplicon**

**Table S16. Cutoff thresholds for hrp2 and hrp3 determination of deletions for each amplicon**.

