## Supplementary files and tables for "Malaria molecular surveillance in the Peruvian Amazon with a novel highly multiplexed *Plasmodium falciparum* Ampliseq assay"

### Supplementary tables and figures

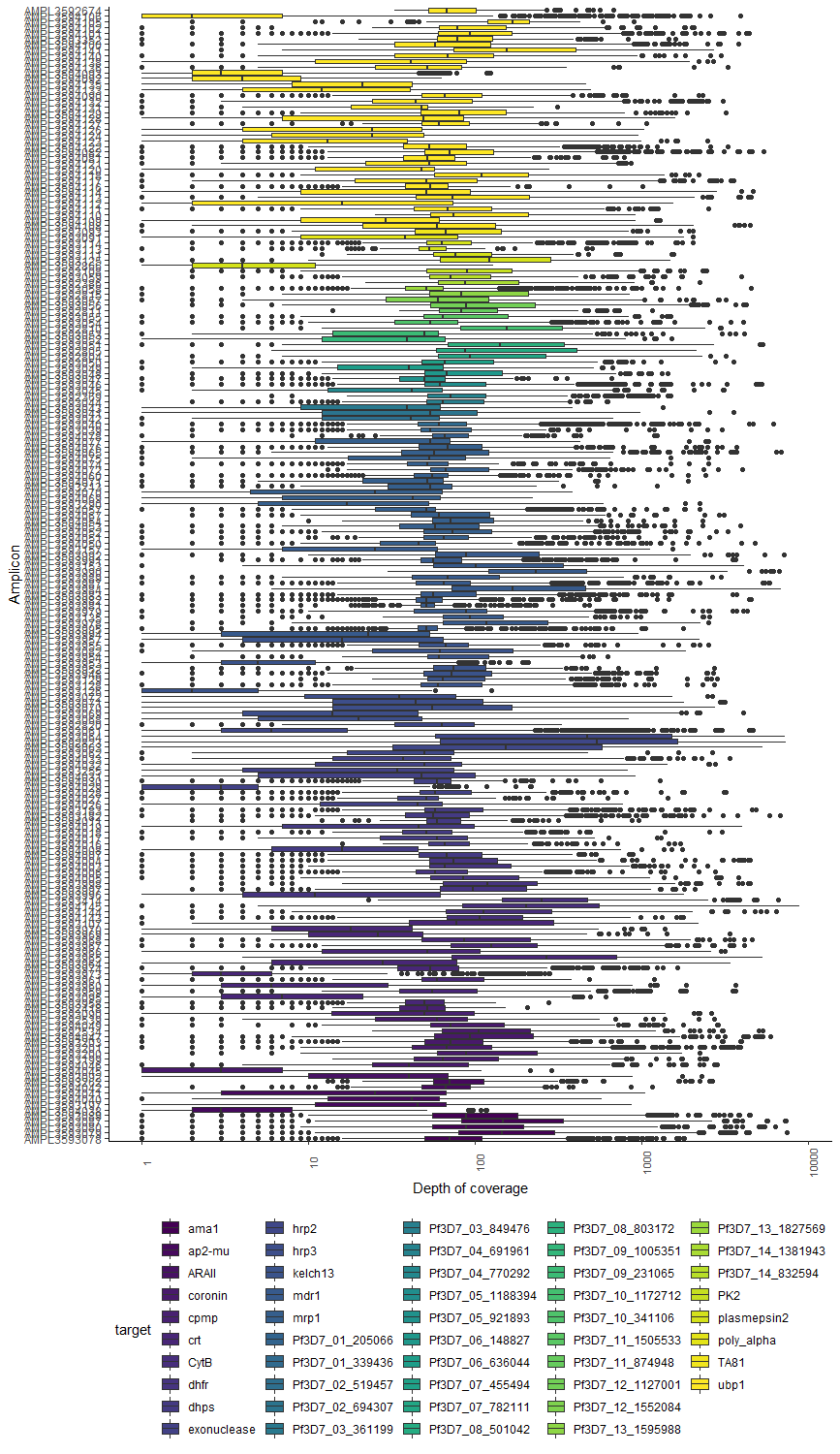

**Figure S1. Distribution of depth of coverage of aligned high quality reads past filter (format field DP) per amplicon region.**

**Table S1.** **Amplicons with low genotype depth.** The proportion of samples or controls with depth of coverage below 10 are listed. When both the controls and the samples have a high proportion of libraries with low depth, then the amplicon is not working well. When there is a higher proportion of samples than controls with low depth, then it is more likely due to variability in the sequence in the study samples. The *hrp2* and *hrp3* amplicons have lower mean depth in the samples than in the controls due to high prevalence of gene deletions in the samples from Peru. There is one *ubp1* amplicon (ubp1_29) that also has poorer performance in samples than controls, possibly due to variations in primer regions in the study samples.

| **Amplicon_ID** | **CHR** | **START** | **END** | **POOL** | **Target** | **Amplicon name** | **% Samples with mean depth <10** | **% controls with mean depth <10** |
| --- | --- | --- | --- | --- | --- | --- | --- | --- |
| AMPL3594045 | Pf3D7_12_v3 | 2091952 | 2092317 | 1 | coronin | coronin_1 | 93% | 100% |
| AMPL3593126 | Pf3D7_13_v3 | 1724575 | 1724896 | 1 | K13 | K13_1 | 94% | 99% |
| AMPL3594029 | Pf3D7_13_v3 | 2503988 | 2504341 | 2 | exonuclease | exonuclease_4 | 94% | 99% |
| AMPL3594038 | Pf3D7_12_v3 | 717766 | 718035 | 1 | ap2-mu | ap2-mu_1 | 91% | 93% |
| AMPL3594106 | Pf3D7_01_v3 | 200597 | 200953 | 1 | ubp1 | ubp1_43 | 95% | 93% |
| AMPL3593973 | Pf3D7_07_v3 | 404305 | 404638 | 1 | crt | crt_7 | 93% | 92% |
| AMPL3594093 | Pf3D7_01_v3 | 197290 | 197637 | 1 | ubp1 | ubp1_29 | 91% | 89% |
| AMPL3592866 | Pf3D7_14_v3 | 293335 | 293566 | 2 | plasmepsin 2 | plasmepsin2_1 | 83% | 85% |
| AMPL3594092 | Pf3D7_01_v3 | 197119 | 197440 | 2 | ubp1 | ubp1_28 | 86% | 85% |
| AMPL3593953 | Pf3D7_13_v3 | 1725868 | 1726167 | 1 | K13 | K13_7 | 78% | 77% |
| AMPL3593061 | Pf3D7_08_v3 | 1374965 | 1375281 | 1 | hrp2 | hrp2_5 | 83% | 72% |
| AMPL3594112 | Pf3D7_01_v3 | 190943 | 191243 | 1 | ubp1 | ubp1_5 | 76% | 69% |
| AMPL3593072 | Pf3D7_13_v3 | 2841623 | 2841818 | 1 | hrp3 | hrp3_5 | 84% | 55% |
| AMPL3593068 | Pf3D7_13_v3 | 2840615 | 2840970 | 1 | hrp3 | hrp3_1 | 82% | 45% |
| AMPL3593069 | Pf3D7_13_v3 | 2840910 | 2841262 | 2 | hrp3 | hrp3_2 | 72% | 35% |

**Table S2.** **Amplicons with high genotype depth of coverage (>150).**

| **Amplicon_ID** | **CHR** | **START** | **END** | **POOL** | **Target** | **Amplicon name** | **median DP** | **25% percentile** | **75% percentile** |
| --- | --- | --- | --- | --- | --- | --- | --- | --- | --- |
| AMPL3592823 | Pf3D7_08_v3 | 1374462 | 1374730 | 1 | hrp2 | hrp2_3 | 268 | 15 | 1045 |
| AMPL3593064 | Pf3D7_08_v3 | 1374687 | 1375025 | 2 | hrp2 | hrp2_4 | 207 | 9 | 905 |
| AMPL3593414 | Pf_M76611 | 4421 | 4659 | 1 | CytB | CytB_5 | 183 | 63 | 329.5 |
| AMPL3593965 | Pf3D7_07_v3 | 404974 | 405187 | 2 | crt | crt_10 | 176 | 59 | 626 |
| AMPL3593990 | Pf3D7_05_v3 | 960959 | 961225 | 2 | mdr1 | mdr1_14 | 155 | 79 | 344 |
| AMPL3594105 | Pf3D7_01_v3 | 200424 | 200657 | 2 | ubp1 | ubp1_42 | 161 | 89.5 | 208 |

**Table S3. Amplicons in conserved regions.** These amplicons had no variants (i.e. only reference sequence detected) in the vcf, and in most cases have few variants detected in South America or even global (source: Pf4 - P. falciparum Community Project Data - Variant catalogue, <https://www.malariagen.net/apps/pf/4.0/#variation>). Only chromosomal variants are included in the Pf4 data-app.

| **Amplicon_ID** | **CHR** | **START** | **END** | **Target** | **Amplicon name** | **nr. variants in Pf4** | **nr. variants with NRAF>0 in SAM Pf4** |
| --- | --- | --- | --- | --- | --- | --- | --- |
| AMPL3594061 | Pf3D7_01_v3 | 467940 | 468211 | mrp1 | mrp1_14 | 9 | 1 |
| AMPL3594073 | Pf3D7_01_v3 | 468287 | 468636 | mrp1 | mrp1_16 | 5 | 0 |
| AMPL3594094 | Pf3D7_01_v3 | 197577 | 197841 | ubp1 | ubp1_30 | 22 | 1 |
| AMPL3594103 | Pf3D7_01_v3 | 199990 | 200337 | ubp1 | ubp1_41 | 22 | 1 |
| AMPL3594125 | Pf3D7_01_v3 | 194496 | 194777 | ubp1 | ubp1_18 | 39 | 0 |
| AMPL3594137 | Pf3D7_01_v3 | 198077 | 198428 | ubp1 | ubp1_32 | 38 | 1 |
| AMPL3594139 | Pf3D7_01_v3 | 198618 | 198838 | ubp1 | ubp1_34 | 11 | 0 |
| AMPL3593191 | Pf3D7_04_v3 | 749304 | 749551 | dhfr | dhfr_7 | 0 | 0 |
| AMPL3593194 | Pf3D7_04_v3 | 749721 | 749960 | dhfr | dhfr_9 | 0 | 0 |
| AMPL3593986 | Pf3D7_05_v3 | 960142 | 960478 | mdr1 | mdr1_10 | 24 | 0 |
| AMPL3593988 | Pf3D7_05_v3 | 960549 | 960738 | mdr1 | mdr1_12 | 20 | 0 |
| AMPL3593972 | Pf3D7_07_v3 | 404272 | 404376 | crt | crt_6 | 3 | 0 |
| AMPL3594014 | Pf3D7_08_v3 | 550217 | 550357 | dhps | dhps_9 | 3 | 0 |
| AMPL3593202 | Pf3D7_12_v3 | 2093070 | 2093321 | coronin | coronin_6 | 10 | 0 |
| AMPL3594039 | Pf3D7_12_v3 | 717975 | 718318 | ap2-mu | ap2-mu_2 | 10 | 0 |
| AMPL3594043 | Pf3D7_12_v3 | 719101 | 719460 | ap2-mu | ap2-mu_7 | 14 | 0 |
| AMPL3594044 | Pf3D7_12_v3 | 719401 | 719727 | ap2-mu | ap2-mu_8 | 12 | 1 |
| AMPL3592420 | Pf3D7_13_v3 | 2503916 | 2504048 | exonuclease | exonuclease_3 | 13 | 0 |
| AMPL3592435 | Pf3D7_13_v3 | 2504281 | 2504427 | exonuclease | exonuclease_5 | 16 | 0 |
| AMPL3593127 | Pf3D7_13_v3 | 1724836 | 1725019 | K13 | K13_2 | 13 | 0 |
| AMPL3593956 | Pf3D7_13_v3 | 1726758 | 1726897 | K13 | K13_11 | 7 | 0 |
| AMPL3593110 | Pf3D7_14_v3 | 293605 | 293961 | plasmepsin2 | plasmepsin2_3 | 23 | 0 |
| AMPL3593186 | Pf_M76611 | 4210 | 4483 | CytB | CytB_4 | NA | NA |

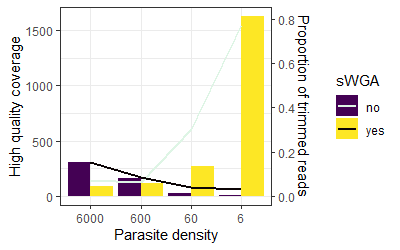

**Figure S2. Effect of selective whole genome amplification (sWGA)** on high quality coverage and amount of trimming of a 3D7 serial dilution at different parasite densities (6000 - 6 p/µl) at DNA concentrations mimicking DBS samples. At parasite densities below 60 p/µl sWGA increases the number of high-quality reads and reduces the number of low-quality reads that are trimmed away.

**Table S4. Error rates in 3D7 replicates without and with sWGA in different subsets of loci.**

| Type of variants | 3D7 without sWGA | 3D7 with sWGA |
| --- | --- | --- |
| All variants in entire target region | 0.05% ± 0.01 | 0.13% ± 0.06 |
| Bi-allelic SNPs only | 0.008% ± 0.004 | 0.03% ± 0.01 |
| Indels only | 0.02% ± 0.005 | 0.04% ± 0.02 |
| “core” region only | 0.03% ± 0.01 | 0.11% ± 0.06 |
| Bi-allelic SNPs in core region only | 0.006% ± 0.004 | 0.02% ± 0.01 |

**Table S5. Genotyping known variants in previously genotyped controls**: MRA 1241, MRA 1251, MRA 1255, MRA 150 (genotypes from literature (72, 92, 102, 103) and samples from Vietnam (104). Several replicates of each samples were tested. NA = no genotype was obtained at this position; wt = wildtype.

| **Sample name** | **PF Ampliseq results** | | | | | **Previous data** | | | | | **error** | **additional mixed** | **total calls ampliseq** | **% error** |
| --- | --- | --- | --- | --- | --- | --- | --- | --- | --- | --- | --- | --- | --- | --- |
|  | ***Crt:* 72-76** | ***K13*** | ***mdr1*: 86, 184, 1246** | ***dhfr:*  51, 59, 108, 164** | ***dhps:* 436, 437, 540, 581, 613** | ***crt:* 72-76** | ***K13*** | ***mdr1:* 86, 184, 1246** | ***dhfr*: 51, 59, 108, 164** | ***Dhps:* 436, 437, 540, 581, 613** |  |  |  |  |
| MRA 150 | CVIET | wt | YYD | IRNI | F,G,K,A,S | CVIET | wt | YYD | IRNI | FGKAS | 0 | 0 | 18 | 0.0% |
|  | NA | wt | YYD | IRNI | F,_,K,A,S |  |  |  |  |  | 0 | 0 | 12 | 0.0% |
|  | CVIET | wt | NA | NA | NA |  |  |  |  |  | 0 | 0 | 6 | 0.0% |
| MRA 1241 | NA | I543T | NYD | IRNL | S/F,G/A,E,A,S | CVIET | I543T | NYD | IRNL | FAEAS | 0 | 2 | 13 | 15.4% |
|  | NA | I543T | N_D | __NI/__NL | _,_,E,A,S |  |  |  |  |  | 0 | 1 | 8 | 12.5% |
|  | CVIET | I543T | N_ | IR__ | S/F,G/A,E,A,A/S |  |  |  |  |  | 0 | 3 | 15 | 20.0% |
| MRA 1251 | CVIET | C580Y | NYD | IRNI | A/S,G,E,A,A | CVIET | C580Y | NYD | IRNI | AGEAA | 0 | 1 | 18 | 5.6% |
|  | NA | NA at pos 580 | N_D | __NI | _,_,E,A,A |  |  |  |  |  | 0 | 0 | 7 | 0.0% |
|  | CVIET | NA at pos 580 | N_ | IR__ | NA |  |  |  |  |  | 0 | 0 | 8 | 0.0% |
| MRA 1255 | CVIET | NA at pos 539 | YYD | IRNI | F,G,K,A,S | CVIET | R539T | YYD | IRNI | FGKAS | 0 | 0 | 17 | 0.0% |
|  | NA | wt | YYD | IRNI | _,_,K,A,S |  |  |  |  |  | 1 | 0 | 11 | 9.1% |
|  | CVIET | NA at pos 539 | NA | IR__/NC__ | NA |  |  |  |  |  | 0 | 2 | 7 | 28.6% |
| VTN 1 | CVIET | T511H | NFD | IRNI | A,G,K/E,A,A | CVIET | Y511H | NFD | IRNI | -GKAA | 0 | 1 | 18 | 0.0% |
|  | NA | T511 +K189T | NFD | NCNI | S,G/A,K,_,A |  |  |  |  |  | 3 | 1 | 13 | 23.1% |
| VTN 2 | CVIDT/CVIET | C580Y/I543T | NYD | IRNI/IRNL | A/S/F,G,E,A,A/S | CVIDT | C580Y | NYD | IRNI | AGEAA | 0 | 4 | 19 | 0.0% |
|  | CVIDT/CVIET | het I543T | NYD | IRNI/IRNL | F,G,E,A,A/S |  |  |  |  |  | 1 | 2 | 18 | 5.6% |
| VTN 3 | CVIET | C580Y/I543T | NYD | IRNI/IRNL | A/S/F,G,K/N,A/G,A/S | CVIET | C580Y | _F_ | IRNL | S_NG_ | 1 | 3 | 19 | 5.3% |
| VTN 4 | CVIET | C580Y | NFD | IRNL | S,G,N,G,A | CVIET | C580Y | _F_ | IRNL | S_NG_ | 0 | 0 | 17 | 0.0% |
|  | NA | C580Y | _FD | IR_L | S,G,-,-,- |  |  |  |  |  | 0 | 0 | 8 | 0.0% |
|  |  |  |  |  |  |  |  |  |  |  | **6** | **20** | **216** | **2.8%** |

**Table S6. Detection limit of variant loci in 3D7-Dd2 mock samples**

| **proportion 3D7 DNA** | **proportion Dd2 DNA** | **nr of heterozygote SNP genotypes** | **nr of homozygote reference SNP genotypes** | **Proportion Dd2 alleles detected** |
| --- | --- | --- | --- | --- |
| 50% | 50% | 63 | 4 | 94.0% |
| 80% | 20% | 28 | 36 | 43.8% |
| 95% | 5% | 3 | 65 | 4.4% |
| 99% | 1% | 0 | 68 | 0% |
| 99.5% | 0.5% | 0 | 69 | 0% |

**Table S7. Complexity of infection analyses.**

COI determined with McCOIL algorithms (categorical and proportional) with different subsets of biallelic variants: 1) all biallelic variants; 2) all variants (core variants) excluding *hrp2*, MS regions and mitochondrial and apicoplast variants; and 3) the 28-SNP barcode variants. Because of the large differences observed between the two McCOIL methods, we estimated the proportions of single and multiple clone infections with an additional methods based on the number of heterozygous variants in 1) the 28-SNP barcode, 2) *ama1*, 3) core variants, and 4) MS targeted regions. The mode (most frequent value) from four measurements for single *vs.* multiple clone (heterozygotes in 1) MS, 2) *ama1* and 3) barcode regions and 4) McCOIL proportional barcode) was determined. The highest number of clones estimated with either the McCOIL categorical or proportional algorithm (categorial uses diploid genotype calls, proportional uses allele depths) was two clones (COI =2). However, a considerably larger proportion of single clone infections was predicted with the categorical method, especially when using more than 28 SNPs. Estimates of single clone infections using the heterozygous loci in the 28-SNP barcode and *ama1* were similar to the 28 SNP proportional McCOIL method (83.9%, 85.0% and 83.1% single clone infections, respectively). With the MS alleles a much larger proportion of multiple clone infections was estimated (61.1%).

|  | | COI=1 | COI>1 | % single clone | 95% CI |
| --- | --- | --- | --- | --- | --- |
| McCOIL categorical | all biallelic variants | 208 | 0 | 100% |  |
|  | core variants | 166 | 1 | 99.4% | 99-100% |
|  | 28-SNP barcode variants. | 192 | 2 | 99.0% | 98-100% |
| McCOIL proportional | all biallelic variants | 3 | 64 | 4.5% | 1.5-9.6% |
|  | core variants | 12 | 112 | 9.7% | 6-15% |
|  | 28-SNP barcode variants. | 123 | 25 | 83.1% | 78-89% |
| Heterozygous variants | MS from NGS | 77 | 121 | 38.9% | 32-46% |
|  | 28-SNP barcode | 213 | 41 | 83.9% | 80-88% |
|  | SNPs in AMA1 | 216 | 38 | 85.0% | 81-89% |
|  | SNPs in core (>5 SNPs) | 85 | 169 | 71.3% | 66-77% |
| **Mode** | | 191 | 51 | 78.9% | 74-84% |

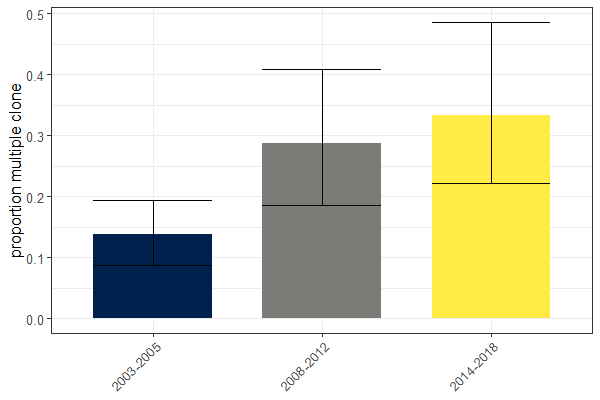

**Figure S3. Proportion multiple clone infections in Peru.** The proportion of multiple clone infections (with 95% confidence interval) was plotted for three time periods, and was higher in 2008-2018 than in 2003-2005 (*p* = 0.0005, Χ^2^). Multiple clone infections determined as mode of the different approaches.

**Table S8. Pairwise comparison of *hrp2/hrp3* classification by PCR and Pf AmpliSeq** in study samples tested with both methods (n = 10). PCR genotypes from Gamboa *et al.* 2010 (2).

| **PCR** | **Pf AmpliSeq** | | | | |
| --- | --- | --- | --- | --- | --- |
|  | *hrp2*+/*hrp3*- | *hrp2*-/*hrp3*- | *hrp2*-/*hrp3*+ | *hrp2* undefined/ *hrp3*+ | **Final result** |
| *hrp2*+/*hrp3*- | 1 |  |  |  | RDT detectable |
| *hrp2*-/*hrp3*- |  | 4 |  |  | RDT failure |
| *hrp2*+/*hrp3*+ |  |  |  | 3 | RDT detectable |
| *hrp2*-/*hrp3*+ |  |  | 1 | 1 | RDT detectable |
| **Final result** | RDT detectable | RDT failure | RDT detectable | RDT detectable |  |

**Table S9 Barcode SNPs**. Chromosomal position and type of the barcode variants are listed. In addition allele frequencies (AF) for the reference allele (REF) and alternate allele (ALT) from PlasmoDB and in study samples (n=254) from Peru.

| CHROM | POS | TYPE | plasmoDB | | | | Samples (n=254) | | |
| --- | --- | --- | --- | --- | --- | --- | --- | --- | --- |
|  |  |  | Reference allele | major allele | Major AF | Minor allele (if not ref) | REF AF | ALT AF | major allele |
| Pf3D7_01_v3 | 205066 | SNP | G | G | 0.66 | A | 0.81 | 0.19 | G |
| Pf3D7_01_v3 | 339436 | SNP | A | G | 0.59 |  | 0.15 | 0.85 | G |
| Pf3D7_02_v3 | 519457 | SNP | C | T | 0.68 |  | 0.61 | 0.39 | C |
| Pf3D7_02_v3 | 694307 | SNP | A | A | 0.54 | G | 0.03 | 0.97 | G |
| Pf3D7_03_v3 | 361199 | SNP | C | C | 0.65 | T | 0.68 | 0.32 | C |
| Pf3D7_03_v3 | 849476 | SNP | C | C | 0.64 |  | 0.99 | 0.01 | C |
| Pf3D7_04_v3 | 691961 | SNP | C | T | 0.53 | T | 0.57 | 0.43 | C |
| Pf3D7_04_v3 | 770292 | SNP | A | A | 0.74 | G | 0.77 | 0.23 | A |
| Pf3D7_05_v3 | 921893 | SNP | T | T | 0.58 | A | 1.00 | 0.00 | T |
| Pf3D7_05_v3 | 1188394 | SNP | C | C | 0.84 | T | 0.79 | 0.21 | C |
| Pf3D7_06_v3 | 148827 | SNP | T | T | 0.8 | C | 0.64 | 0.36 | T |
| Pf3D7_06_v3 | 636044 | SNP | A | C | 0.58 |  | 0.67 | 0.33 | A |
| Pf3D7_07_v3 | 455494 | SNP | A | A | 0.57 |  | 0.79 | 0.21 | A |
| Pf3D7_07_v3 | 782111 | SNP | T | T | 0.92 | G | 0.91 | 0.09 | T |
| Pf3D7_08_v3 | 501042 | SNP | T | T | 0.56 | C | 0.27 | 0.73 | C |
| Pf3D7_08_v3 | 803172 | SNP | T | T | 0.82 | G | 0.10 | 0.90 | G |
| Pf3D7_09_v3 | 231065 | SNP | C | A | 0.5 |  | 0.80 | 0.20 | C |
| Pf3D7_09_v3 | 1005351 | SNP | G | G | 0.65 | C | 0.49 | 0.51 | C |
| Pf3D7_10_v3 | 341106 | SNP | A | A | 0.54 | G | 0.01 | 0.99 | G |
| Pf3D7_10_v3 | 1172712 | SNP | A | G | 0.65 | G | 0.25 | 0.75 | G |
| Pf3D7_11_v3 | 874948 | SNP | G | A | 0.7 | A | 0.43 | 0.57 | A |
| Pf3D7_11_v3 | 1505533 | SNP | T | T | 0.92 | C | 0.98 | 0.02 | T |
| Pf3D7_12_v3 | 1127000 | INDEL | T |  |  |  | 0.30 | 0.70 | TA |
| Pf3D7_12_v3 | 1552084 | SNP | T | C | 0.86 | C | 0.29 | 0.71 | C |
| Pf3D7_13_v3 | 1595988 | SNP | T | C | 0.54 | C | 0.68 | 0.32 | T |
| Pf3D7_13_v3 | 1827569 | SNP | T | T | 0.62 | A | 0.50 | 0.50 | T |
| Pf3D7_14_v3 | 832594 | SNP | T | T | 0.87 | C | 0.61 | 0.39 | T |
| Pf3D7_14_v3 | 1381943 | SNP | T | T | 0.94 | C | 0.01 | 0.99 | C |

**Figure S4. Principal component analysis of samples (n=254) collected in Peru between 2003 and 2018**. PCA is shown all biallelic loci in the core region along the first 2 principal components (A) and 3^rd^ and 4^th^ PCs (B). Isolates are colored by year (A& B) and by district (C &D), and from earlier years (blue and purple colors) are more diverse than later isolates (greens & yellows), which form two clusters. All samples with unknown district were collected in the rural communities south of Iquitos.

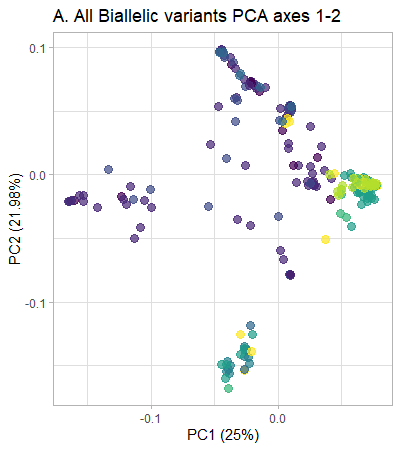

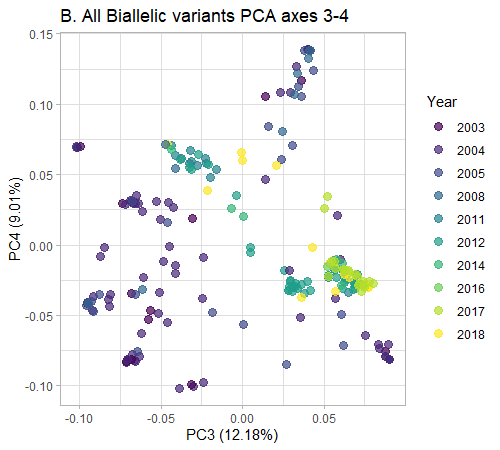

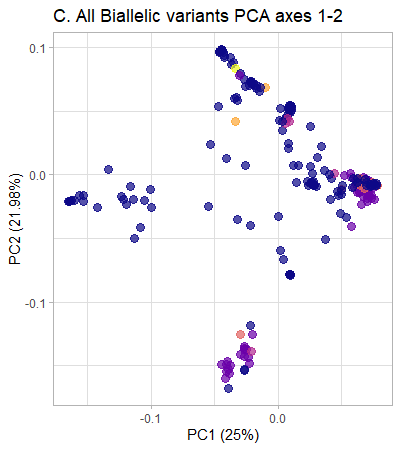

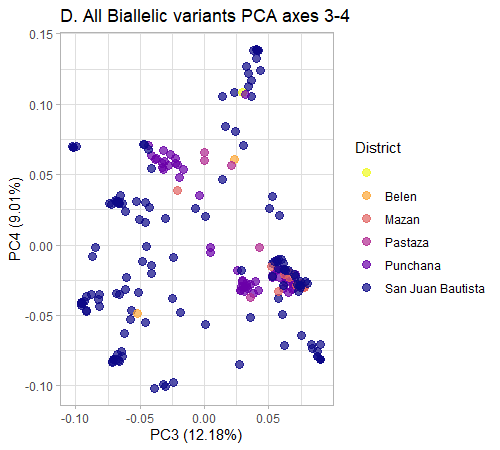

**Figure S5. Expected Heterozygosity by time period and district.** Number of individuals for each population: n_2003-2005_ = 1, n_2003-2005_Belen_ = 1, n_2003-2005_San Juan Bautista_ = 116, n_2008-2012_Punchana_ = 59, n_2008-2012_San Juan Bautista_ = 6, n_2014-2018_San Juan Bautista_ = 24, n_2014-2018_Mazan_ = 10, n_2014-2018_Pastaza_ = 4.

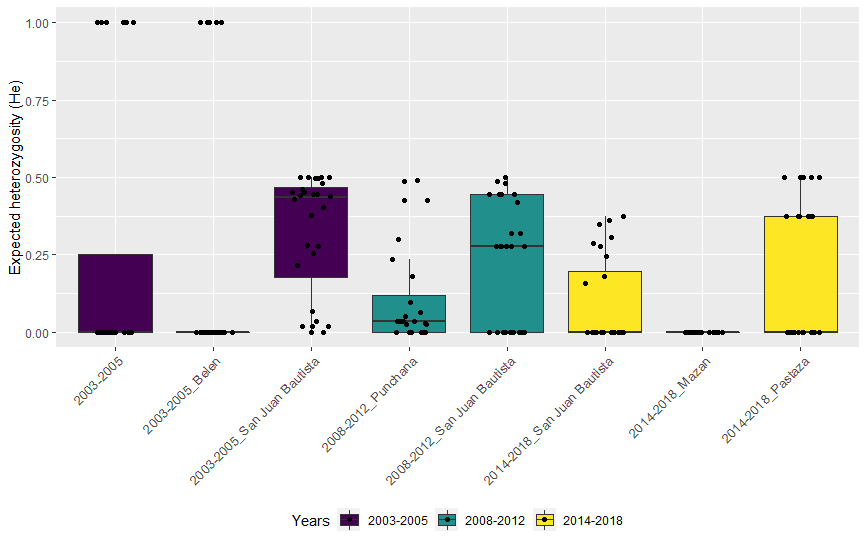

**Table S10. p-values for pairwise comparisons of *He*** using Wilcoxon rank sum test with Benjamini-Hochberg correction for multiple testing. Significant p-values (<0.05) are indicated in bold.

|  | 2003-2005  unspecified | 2003-2005  Belen | 2003-2005 San Juan Bautista | 2008-2012  Punchana | 2008-2012  San Juan Bautista | 2014-2018  San Juan Bautista | 2014-2018  Mazan |
| --- | --- | --- | --- | --- | --- | --- | --- |
| 2003-2005 Belen | 0.61 | - | - | - | - | - | - |
| 2003-2005 San Juan Bautista | **0.009** | **>0.001** | - | - | - | - | - |
| 2008-2012 Punchana | 0.117 | **0.018** | **0.0010** | - | - | - | - |
| 2008-2012 San Juan Bautista | 0.20 | **0.039** | **0.045** | 0.24 | - | - | - |
| 2014-2018 San Juan Bautista | 0.95 | 0.58 | **>0.001** | 0.12 | **0.018** | - | - |
| 2014-2018 Mazan | **0.015** | **0.039** | **>0.001** | **>0.001** | **>0.001** | **0.0046** | - |
| 2014-2018 Pastaza | 0.66 | 0.23 | **0.034** | 0.93 | 0.72 | 0.12 | **>0.001** |

**Figure S6. Genetic differentiation between parasite populations in the three time periods,** measured as Fst (Weir & Cockerham, 1984), G`_ST_ (Hedrick, 2005) and Jost's D (Jost 2008) using the R package diveRsity. Number of individuals for each population: n_2003-2005_ = 118; n_2008-2012_ = 65; n_2014-2018_ = 38.

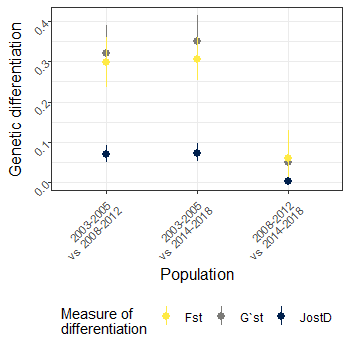

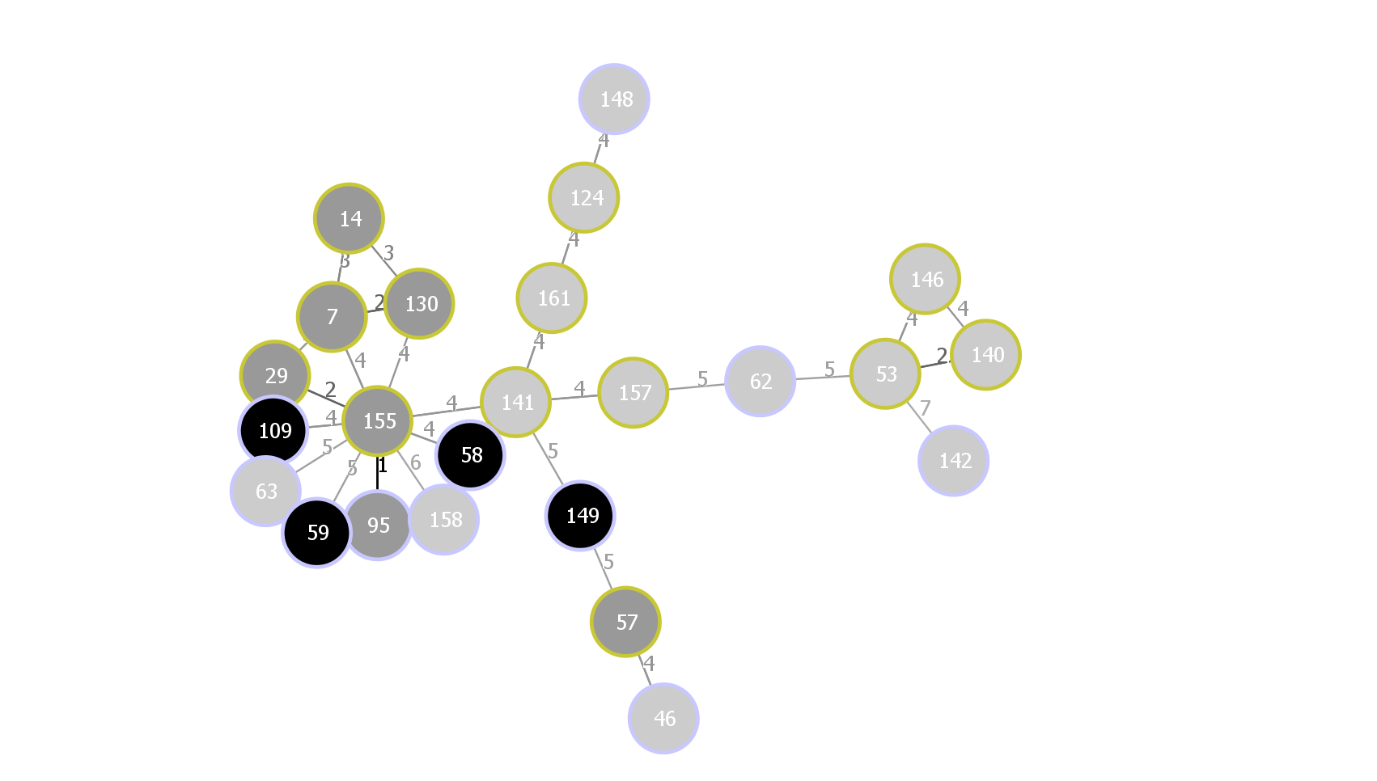

**Figure S7. Minimum spanning network (nLV graph) of multilocus lineages.** Minimum spanning network generated with goeBURST distance, nLV =4, created in Phyloviz v2.0 with barcode genotypes for each lineage. Number correspond to lineages from table 2 main document. Light grey: first observed in 2003-2005, dark-grey: first observed in 2008-2012, black: first observed in 2014-2018. After 2008, lineage no. 155 becomes predominant, with many other circulating lineages in that time related to lineage 155. Lineage 149 and 57 found in Pastaza in 2018 had a different origin.

**Table S11.** **Linkage disequilibrium** expressed as $\bar{r}D$ per population (time period and/or district), measured with 999 resamplings using the poppr package in R.

| years | district | n | $\bar{r}D$ | p-value |
| --- | --- | --- | --- | --- |
| 2003-2018 | All | 221 | 0.155 | 0.001 |
| 2003-2005 | All | 118 | 0.149 | 0.001 |
| 2008-2012 | All | 65 | 0.250 | 0.001 |
| 2014-2018 | All | 38 | 0.299 | 0.001 |
| 2003-2005 | San Juan Bautista | 116 | 0.15 | 0.001 |
| 2008-2012 | San Juan Bautista | 6 | 0.38 | 0.001 |
| 2008_2012 | Punchana | 59 | 0.222 | 0.001 |
| 2014-2018 | San Juan Bautista | 24 | 0.408 | 0.001 |

**Table S12. 28-SNP barcode loci that become fixed over time**

| Chromosome | Position | Fixed in |
| --- | --- | --- |
| Pf3D7_02_v3 | 519457 | 2014-2018 |
| Pf3D7_02_v3 | 694307 | 2014-2018 |
| Pf3D7_03_v3 | 849476 | 2008-2018 |
| Pf3D7_04_v3 | 691961 | 2014-2018 |
| Pf3D7_05_v3 | 921893 | 2003-2018 |
| Pf3D7_06_v3 | 636044 | 2014-2018 |
| Pf3D7_07_v3 | 455494 | 2014-2018 |
| Pf3D7_07_v3 | 782111 | 2008-2018 |
| Pf3D7_08_v3 | 803172 | 2008-2018 |
| Pf3D7_10_v3 | 341106 | 2003-2018 |
| Pf3D7_11_v3 | 1505533 | 2008-2018 |
| Pf3D7_12_v3 | 1127000 | 2014-2018 |
| Pf3D7_12_v3 | 1552084 | 2008-2018 |
| Pf3D7_14_v3 | 1381943 | 2008-2018 |

**Table S13 Contributions of alleles to DAPC.**

| **variant position** | **contributing factor** | **PC in DAPC** | **annotation of alt allele** | **gene ID** | **gene name** | **28-SNP barcode** |
| --- | --- | --- | --- | --- | --- | --- |
| Pf3D7_01_v3_192590 | 0.013224389 | 2 | Lys774Asn | PF3D7_0104300 | ubp1 |  |
| Pf3D7_01_v3_196974 | 0.011249826 | 2 | Leu2236Leu | PF3D7_0104300 | ubp1 |  |
| Pf3D7_01_v3_199237 | 0.008442663 | 2 | Lys2912Asn | PF3D7_0104300 | ubp1 |  |
| Pf3D7_01_v3_205066 | 0.007943108 | 2 | Phe130Phe | PF3D7_0104500 | unknown protein | Yes |
| Pf3D7_01_v3_339432 | 0.035879398 | 1 | Glu755Glu | PF3D7_0108300 | unknown protein | in barcode amplicon |
| Pf3D7_02_v3_519457 | 0.025872597 | 2 | Ile3228Ile | PF3D7_0212500 | unknown protein | in barcode amplicon |
| Pf3D7_03_v3_361195 | 0.007454305 | 2 | Arg1626Lys | PF3D7_0308100 | zinc finger protein, putative | in barcode amplicon |
| Pf3D7_03_v3_361199 | 0.008475745 | 2 | Thr1627Thr | PF3D7_0308100 | zinc finger protein, putative | Yes |
| Pf3D7_04_v3_748235 | 0.0860365 | 1 | Cys50Arg | PF3D7_0417200 | pfdhfr |  |
| Pf3D7_04_v3_748235 | 0.009881925 | 2 | Cys50Arg | PF3D7_0417200 | pfdhfr |  |
| Pf3D7_04_v3_748239 | 0.01967936 | 1 | Asn51Ile | PF3D7_0417200 | pfdhfr |  |
| Pf3D7_04_v3_748239 | 0.009615098 | 2 | Asn51Ile | PF3D7_0417200 | pfdhfr |  |
| Pf3D7_04_v3_748577 | 0.02383201 | 1 | Ile164Leu | PF3D7_0417200 | pfdhfr |  |
| Pf3D7_04_v3_770292 | 0.032732908 | 2 | Lys4667Glu | PF3D7_0417400 | unknown protein | Yes |
| Pf3D7_05_v3_1188491 | 0.032529119 | 1 | Phe152Phe | PF3D7_0529000 | unknown protein | in barcode amplicon |
| Pf3D7_05_v3_960989 | 0.01175819 | 1 | Ser1034Cys | PF3D7_0523000 | pfmdr1 |  |
| Pf3D7_05_v3_960989 | 0.009963482 | 2 | Ser1034Cys | PF3D7_0523000 | pfmdr1 |  |
| Pf3D7_05_v3_961625 | 0.031696855 | 1 | Asp1246Tyr | PF3D7_0523000 | pfmdr1 |  |
| Pf3D7_06_v3_148827 | 0.010701516 | 1 | Val39Val | PF3D7_0603600 | AT-rich interactive domain-containing protein, putative | Yes |
| Pf3D7_06_v3_148827 | 0.017232716 | 2 | Val39Val | PF3D7_0603600 | AT-rich interactive domain-containing protein, putative | Yes |
| Pf3D7_06_v3_636044 | 0.046900879 | 2 | Ile1310Met | PF3D7_0615400 | ribonuclease, putative | Yes |
| Pf3D7_07_v3_455494 | 0.007594591 | 2 | Asn927Asn | PF3D7_0710100 | unknown protein | Yes |
| Pf3D7_07_v3_455550 | 0.007107321 | 2 | Ile909Leu | PF3D7_0710100 | unknown protein | in barcode amplicon |
| Pf3D7_08_v3_501042 | 0.018331353 | 2 | Glu176Glu | PF3D7_0809700 | RuvB-like helicase 1 | Yes |
| Pf3D7_08_v3_501054 | 0.017953778 | 2 | Val172Val | PF3D7_0809700 | RuvB-like helicase 1 | in barcode amplicon |
| Pf3D7_08_v3_549993 | 0.024643639 | 1 | Lys540Glu | PF3D7_0810800 | pfdhps |  |
| Pf3D7_08_v3_549993 | 0.007753441 | 2 | Lys540Glu | PF3D7_0810800 | pfdhps |  |
| Pf3D7_09_v3_1005351 | 0.036558277 | 2 | Glu1919Gln | PF3D7_0924600 | unknown protein | Yes |
| Pf3D7_11_v3_874948 | 0.012525102 | 2 | Gly973Asp | PF3D7_1122800 | calcium-dependent protein kinase 6 | Yes |
| Pf3D7_12_v3_2092606 | 0.013989583 | 2 | Val62Met | PF3D7_1251200 | pfcoronin |  |
| Pf3D7_12_v3_2093692 | 0.014452113 | 2 | Val424Ile | PF3D7_1251200 | pfcoronin |  |
| Pf3D7_12_v3_2094242 | 0.00941445 | 1 | downstream variant | PF3D7_1251200 | pfcoronin |  |
| Pf3D7_13_v3_1827569 | 0.016409733 | 1 | Tyr345Asn | PF3D7_1345600 | inner membrane complex protein | Yes |
| Pf3D7_13_v3_1827569 | 0.010051953 | 2 | Tyr345Asn | PF3D7_1345600 | inner membrane complex protein | Yes |
| Pf3D7_14_v3_294796 | 0.022437109 | 2 | Gln442His | PF3D7_1408000 | plasmepsin II |  |

**Figure S8. Copy number variations in A) *plasmepsin II* gene (*pm2*) and B) *multidrug resistance gene 1* (*mdr1*)** in a subset of samples from Peru collected between 2003-2018. Samples with copy numbers between 0.5 - 1.5 (dotted lines) relative to 3D7 are considered to have single copies of the respective genes. Sample sizes: n_2003-2005_ = 31, n_2008-2012_ = 13, n_2014-2018_ = 34.

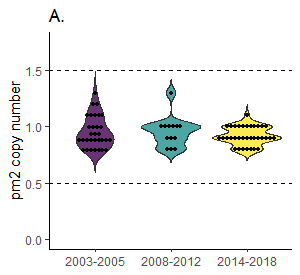

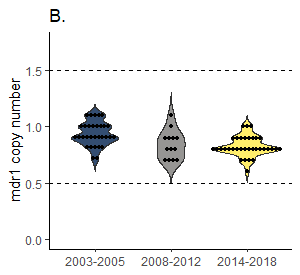

**Table S14 Cost comparison AmpliSeq vs WGS at comparable depth and per kit.**

| **WGS on MiSeq** | **Price for 1 kit (24 samples)** | |  |  | **Price per sample for 1 run with 12 samples (20X-50X coverage)** | |
| --- | --- | --- | --- | --- | --- | --- |
|  | **Peru** | **Belgium** |  |  | **Peru** | **Belgium** |
| Nextera XT DNA Library Kit (24 samples) | $1,520 | $960 |  |  | $63 | $40 |
| Nextera XT Index Kit (24 indexes) | $455 | $290 |  |  | $19 | $12 |
| MiSeq Reagent Kit v3 (600-cycles) | $2,415 | $1,780 |  |  | $201 | $148 |
| **total** | $4,390 | $3,030 |  |  | **$283** | **$200** |
| **AmpliSeq on Miseq** | **Price for 1 kit (96 samples)** | | **Price per sample for 1 run with 96 samples (500X-1000X coverage)** | | **Price per sample for 1 run with 384 samples (50X-100X coverage)** | |
|  | **Peru** | **Belgium** | **Peru** | **Belgium** | **Peru** | **Belgium** |
| Ampliseq library plus for 96 samples | $14,445 | $10,840 | $150 | $113 | $150 | $113 |
| AmpliSeq Index kit | $950 | $715 | $10 | $7 | $10 | $7 |
| Miseq reagent kit v3 | $2,415 | $1,780 | $25 | $19 | $6 | $5 |
| custom pools | $2,210 | $2,210 | $0.37 | $0.37 | $0.37 | $0.37 |
| **total** | $20,020 | $15,545 | $185 | $139 | **$166** | **$125** |
| Note: these prices are in USD and rounded for the table. Belgian prices in euros have been converted to USD with an exchange rate of 0.88 USD to 1 EUR. | | | | | | |
| Prices for WGS analysis do not include preprocessing of the samples, for example with sWGA, which is usually required for DBS samples and would add another $30-$50 per sample | | | | | | |
| Secondary reagents required during both library preparation procedures not included in the kits, such as AMPureXP beads are not included in these prices and add similar costs to both. | | | | | | |

**Table S15: Laboratory strains included in assay validation**

| **Laboratory isolates** | tested with sWGA |
| --- | --- |
| 3D7 |  |
| Dd2 (MRA-150) |  |
| CamWT_C580Y (MRA-1251) |  |
| Dd2_R539T (MRA-1255) |  |
| IPC 4912 (MRA-124) |  |
| ten-fold serial dilution of 3D7 parasite DNA (60.000 p/µl to 6 p/µl) | x |
| 3D7:Dd2 mixtures (20,000 p/µL): 50-50% ratio, 80-20%, 95-5%, 99-1% and 99.5-0.5% |  |
| Uninfected human DNA |  |

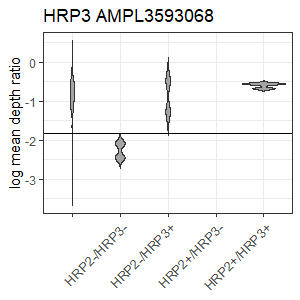

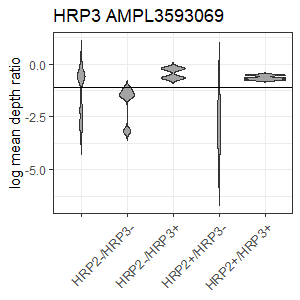

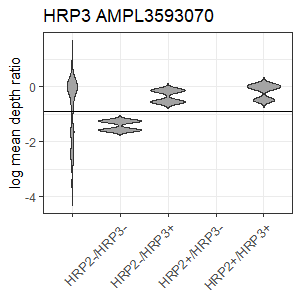

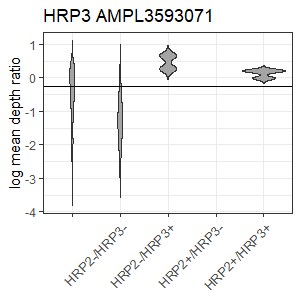

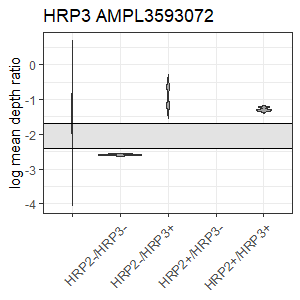

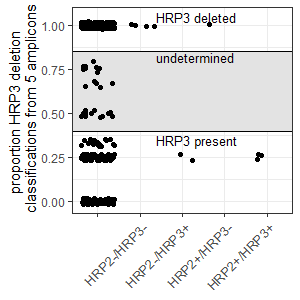

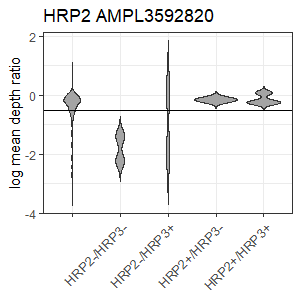

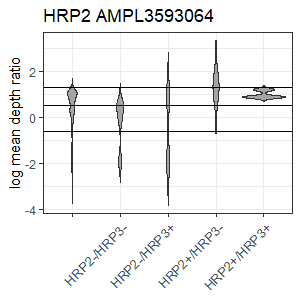

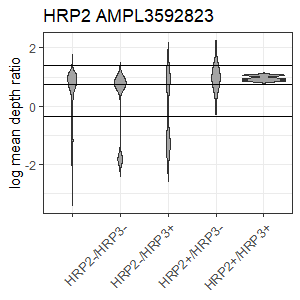

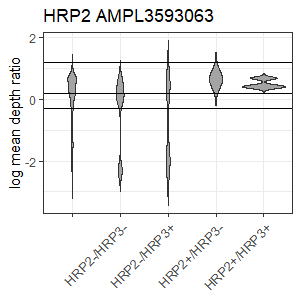

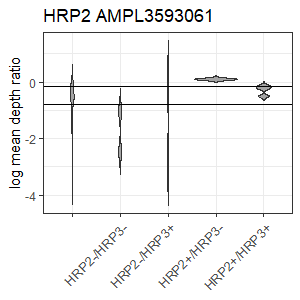

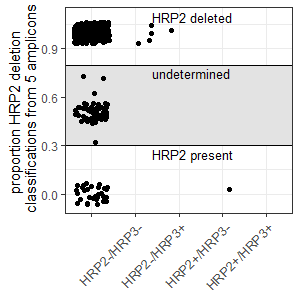

**Figure S9. Distributions of log mean depth ratio’s for all samples for each *hrp3* amplicon (AMPL3593072, AMPL3593071, AMPL3593070, AMPL3593069, AMPL3593068) and *hrp2* amplicon** (AMPL3592820, AMPL3593064, AMPL3592823, AMPL3593063, AMPL3593061), plotted by *hrp2*/*hrp3* PCR results (not tested, *hrp2*-/*hrp3*-, *hrp2*-/*hrp3*+, *hrp2*+/*hrp3*-, *hrp2*+/*hrp3*+), with thresholds used to define deletions or presence of the genes (Supp. Table 2). To classify a sample as *hrp2* and *hrp3* deleted or non-deleted, the number of amplicons per sample and gene with deletions was summed and then divided by the total number of amplicons (with or without the deletion). If the resulting ratio was >0.8 a sample was classified as having a deletion in *hrp3* or *hrp2*; if the ratio was < 0.3 for hrp2 or <0.4 for hrp3, the samples was classified as without deletion in that gene.

**Table S16. Cutoff thresholds for *hrp2* and *hrp3* determination of deletions for each amplicon.**

| **Gene** | **Amplicon** | **Threshold log mean depth ratio** | **Result** | |
| --- | --- | --- | --- | --- |
| *hrp3* | AMPL3593072 | <= -2.4 | *hrp3*- | |
|  | AMPL3593072 | >= -1.7 | *hrp3*+ | |
|  | AMPL3593071 | <= -0.25 | *hrp3*- | |
|  | AMPL3593070 | < -0.9 | *hrp3*- | |
|  | AMPL3593070 | >= -0.9 | *hrp3*+ | |
|  | AMPL3593069 | < -1.10 | *hrp3*- | |
|  | AMPL3593068 | < -1.82 | *hrp3*- | |
|  | AMPL3593068 | >= -1.82 | *hrp3*+ | |
| *hrp2* | AMPL3592820 | < -0.50 | *hrp2*- | |
|  | AMPL3592820 | >= -0.50 | & *hrp3*- | *hrp2*+ |
|  |  |  | & *hrp3*+ | undetermined |
|  | AMPL3593064 | < -0.60 | *hrp2*- | |
|  | AMPL3593064 | < 0.5 | & *hrp3*+ | *hrp2*- |
|  | AMPL3593064 | >1.30 | & *hrp3*- | *hrp2*+ |
|  | AMPL3592823 | <-0.35 | *hrp2*- | |
|  | AMPL3592823 | <0.75 | & *hrp3*+ | *hrp2*- |
|  | AMPL3592823 | >1.40 | & *hrp3*- | *hrp2*+ |
|  | AMPL3593063 | < -0.3 | *hrp2*- | |
|  | AMPL3593063 | < 0.2 | & *hrp3*+ | *hrp2*- |
|  | AMPL3593063 | >1.2 | & *hrp3*- | *hrp2*+ |
|  | AMPL3593061 | < -0.15 | & *hrp3*- | *hrp2*- |
|  | AMPL3593061 | >= -0.15 | & *hrp3*- | *hrp2*+ |
|  | AMPL3593061 | < -0.8 | & *hrp3*+ | *hrp2*- |
|  | AMPL3593062 | Not used; no discriminatory power | | |
